# The impact of varying the number and selection of conditions on estimated multimorbidity prevalence: a cross-sectional study using a large, primary care population dataset

**DOI:** 10.1101/2023.02.16.23285983

**Authors:** Clare MacRae, Megan McMinn, Stewart W Mercer, David Henderson, David A McAllister, Iris Ho, Emily Jefferson, Daniel R Morales, Jane Lyons, Ronan A Lyons, Chris Dibben, Bruce Guthrie

## Abstract

**Background:** Multimorbidity prevalence rates vary considerably depending on the conditions considered in the morbidity count, but there is no standardised approach to the number or selection of conditions to include.

**Methods and Findings:** We conducted a cross-sectional study using English primary care data for 1168260 participants who were all people alive and permanently registered with 149 included general practices. Outcome measures of the study were prevalence estimates of multimorbidity when varying the number and selection of conditions considered (≥two conditions) for 80 conditions. Included conditions featured in ≥one of the nine published lists of conditions examined in the study and/or phenotyping algorithms in the Health Data Research UK Phenotype Library. First, multimorbidity prevalence was calculated when considering the individually most common two conditions, three conditions, etc, up to 80 conditions. Second, prevalence was calculated using nine condition-lists from published studies. Analyses were stratified by dependent variables age, socioeconomic position, and sex. Prevalence when only the two commonest conditions were considered was 4.6% (95%CI [4.6,4.6] p <0.001), rising to 29.5% (95%CI [29.5,29.6] p <0.001) considering the 10 commonest, 35.2% (95%CI [35.1,35.3] p <0.001) considering the 20 commonest, and 40.5% (95%CI [40.4,40.6] p <0.001) when considering all 80 conditions. The threshold number of conditions at which multimorbidity prevalence was >99% of that measured when considering all 80 conditions was 52 for the whole population but was lower in older people (29 in >80 years) and higher in younger people (71 in 0–9-year-olds). Nine published condition-lists were examined; these were either recommended for measuring multimorbidity, used in previous highly cited studies of multimorbidity prevalence, or widely applied measures of ‘comorbidity’. Multimorbidity prevalence using these lists varied from 11.1% to 36.4%. A limitation of the study is that conditions were not always replicated using the same ascertainment rules as previous studies to improve comparability across condition lists, but this highlights further variability in prevalence estimates across studies.

**Conclusions:** In this study we observed that varying the number and selection of conditions results in very large differences in multimorbidity prevalence, and different numbers of conditions are needed to reach ceiling rates of multimorbidity prevalence in certain groups of people. These findings imply that there is a need for a standardised approach to defining multimorbidity, and to facilitate this, researchers can use existing condition-lists associated with highest multimorbidity prevalence.

**Author summary:** *Why was this study done?:*

- There is wide variety in the conditions considered by researchers when measuring multimorbidity prevalence.
- A systematic review of 566 studies, published in 2021, found lack of consensus in the selection of conditions considered.
- In half of studies only eight conditions (diabetes, stroke, cancer, chronic obstructive pulmonary disease, hypertension, coronary heart disease, chronic kidney disease, and heart failure) were consistently considered; and the number of conditions considered varied from 2 to 285 (median 17).
- A more consistent approach to measuring multimorbidity is needed to facilitate comparability and generalisability across studies.

*What did the researchers do and find?:*

- This study investigated the relationship between the number and selection of conditions considered and the impact on multimorbidity prevalence.
- There are large differences in prevalence, a range of 4.6% to 40.5%, when different numbers and selections of conditions are considered.
- Nine published condition-lists were examined; including those recommended for measuring multimorbidity, previously used to measure multimorbidity prevalence, or measures of ‘comorbidity’.
- Highest multimorbidity prevalence was found when using Ho always + usually (a list derived from a recent Delphi consensus study), Barnett (widely used to measure multimorbidity prevalence), and Fortin (a list recommended for use in measuring multimorbidity).
- People who are the oldest, living in the most deprived areas, and men require fewer conditions to be considered to reach close to multimorbidity prevalence when considering all 80 conditions (the ceiling effect, where the prevalence approaches the upper limit of prevalence possible in the study).

*What do these findings mean?:*

- All conditions were counted in the same way (the presence of the condition ever recorded) to improve comparability, however in previous studies conditions were counted according to varying rules, highlighting that further variability in prevalence estimates across studies will happen because of variation in how each condition is measured.
- There is a need for standardisation when measuring multimorbidity prevalence so that results across studies are comparable and population subgroups are accurately represented.
- To address this, researchers can consider using the Ho always + usually, Barnett, or Fortin condition lists.

## Introduction

Multimorbidity is increasing in prevalence due to improved survival from chronic diseases and population ageing, and now poses major challenges to healthcare systems worldwide [1]. Multimorbidity is common, increases substantially with advancing age, and is more common in women and people with lower socioeconomic position (SEP) [2] [3]. Despite its importance, existing research literature is highly heterogenous in how it defines and measures multimorbidity [4]. Choice of conditions considered in the count (the denominator) when measuring multimorbidity prevalence is likely to be driven by pragmatic decision making in the context of data availability [5], or by recycling of existing published condition lists, and results in wide diversity in the number and selection of conditions considered in current multimorbidity literature [6]. In a systematic review of 566 studies of multimorbidity, the number of conditions considered in counts of multimorbidity prevalence ranged from two to 285 (median 17, interquartile range [IQR] 11-23) [4]. Only eight core conditions were consistently considered in more than half of studies (diabetes, stroke, cancer, chronic obstructive pulmonary disease, hypertension, coronary heart disease, chronic kidney disease, and heart failure) [4].

As a result of this diversity, multimorbidity prevalence estimates vary widely across studies [7], making it difficult to make comparisons within the existing literature. Unsurprisingly, higher multimorbidity prevalence is reported by studies that consider a larger number of conditions in their count of multimorbidity [7, 8], studies that consider conditions that are most prevalent [2], and in studies that include more people in older age-groups [7]. The number and selection of conditions considered when measuring multimorbidity prevalence is therefore important, but there is little consistent guidance to support researchers when deciding which conditions to consider. Researchers have recommended condition-lists to consider in multimorbidity measurement, including 11 conditions by Diederichs et al [6], and 20 conditions by Fortin et al [9], More recently, a modified Delphi panel study by Ho et al[10] developed two condition-lists based on international consensus on the measurement of multimorbidity: one list recommending conditions to always consider, and a second recommending conditions to usually consider when counting multimorbidity prevalence [10].

All multimorbidity research findings are dependent on decisions made at the earliest stages in measurement, including what has been measured, and therefore building understanding of the properties of multimorbidity as a concept is needed. To inform researchers’ choices, and improve the comparability and reproducibility of future research, it is important to understand the relationship between multimorbidity prevalence and the number and selection of conditions considered in the count. The aim of this study was to examine these relationships in a large primary care cohort.

## Methods

### Study design

A cross-sectional study design was used to examine the hypothesis that multimorbidity prevalence varies when considering different numbers of conditions, and different selections of conditions (using published recommended or commonly used condition-lists), in the count. The analyses were designed in November 2021, performed in February 2022, and no data-driven changes to analyses took place during this period. As part of the peer review process, one additional condition-list was added [11], and the paper updated to include p-values as well as confidence intervals for proportions, and sensitivity analyses of variation in prevalence by deprivation and gender using raw rather than direct age-standardised data were added.

### Data sources

The study analysed routinely collected, anonymised individual level data from English participants in the Clinical Practice Research Datalink (CPRD) Gold dataset, which are broadly representative of the UK population [12]. Available data included individual demography (age, SEP, and sex), clinical codes from both GP electronic health records (Read codes) and hospital admission data (ICD-10 codes), and laboratory results. Socioeconomic position (SEP) was defined as deciles of the English Index of Multiple Deprivation (IMD), a measure of relative deprivation according to small local area level, with deciles defined by national thresholds [13].

### Study participants

Study participants were all people who were alive and permanently registered with 149 included general practices on the study index date, November 30, 2015, with least two year’s GP registration prior to this [14].

### Definition of variables

For each individual, we defined the presence of 80 conditions using a set of existing code-lists which combined Read codes (version 2) applied to GP electronic health record data, International Classification of Diseases 10^th^ version (ICD-10) codes applied to hospital admission data, and laboratory results recorded in the GP electronic health record [15] (Supplementary Table 1). The 80 conditions were chosen because they featured in one or more of the nine published lists of conditions examined in the study and/or phenotyping algorithms (condition code lists) in the HDR-UK Phenotype Library [15]. New code lists were made by study authors for conditions featured in exiting condition-lists where no HDR-UK algorithms were available and are listed in Supplementary Table 1. All the codes used to identify individuals with each condition were mutually exclusive, therefore double counting of conditions was not possible, and all conditions contained within condition-lists were included in the total 80 conditions.

Condition-lists were either specifically recommended for measuring multimorbidity (referred to hereinafter as Diederichs [6], Fortin [9], Ho always [10], Ho always + usually [10], and N’Goran [11]), used in previous highly cited large-scale studies of multimorbidity prevalence (Barnett[16] and Salive [17]), or included in widely applied measures of ‘comorbidity’ (Charlson [18] and Elixhauser [19]). The two condition-lists recommended by the recent Ho et al Delphi consensus study [10] were conditions recommended to always include (Ho always), and all the conditions recommended by both the lists, conditions to always include and to usually include (Ho always + usually) (Supplementary Information Panel).

Heterogeneity existed in the description and the hierarchical level of conditions between condition-lists. Therefore, to ascertain each condition in the same way for every condition-list, some were dis-aggregated to more granular descriptions. For example, Diederichs et al [6] considered cancer, while Ho always [10] considered three condition groups that were all cancers: primary malignancy, secondary malignancy, and haematological malignancy. In this case, in the Diederichs et al [6] condition-list, cancer was inflated from one to three conditions (to primary malignancy, secondary malignancy, and haematological malignancy) to allow direct comparison with Ho always [10]. Therefore, the number of included conditions in some condition-lists varied from the original published lists (Supplementary Table 2).

### Statistical analysis

Multimorbidity prevalence was calculated when different numbers and selections of conditions were considered in the count (the denominator). In all analyses, multimorbidity was defined by the cut-off (the numerator) which was the presence of ≥ two conditions [3].

We conducted a suite of comparisons including examination of the effect of the number of conditions considered in the count on multimorbidity prevalence by considering the most common two conditions, followed by the most common three conditions, the most common four conditions, etc, for every number up to considering all 80 conditions in the count (Information Panel). To do this, conditions were ordered from most to least prevalent and added in turn to each new count. The prop.test procedure in R was used to estimate 95% confidence intervals and ps for prevalence using the normal approximation for large samples.

By making the assumption that multimorbidity prevalence estimated by considering all 80 conditions in the count was the true prevalence, we then calculated the number of conditions that had to be included to exceed a relative risk (RR) of 0.99 of this ‘true’ prevalence. This was done to estimate when a ceiling effect was present, where the prevalence approaches the upper limit of possible prevalence in the study and the point at which adding further conditions to the count had very little impact on multimorbidity prevalence.

To examine the effect the selection of conditions considered in the count, multimorbidity prevalence was calculated when considering the conditions included in each of the nine condition-lists. Since age is very strongly associated with multimorbidity and the SEP and sex composition within different age-groups varies making age a major confounder, analyses were standardised to the age-structure of the whole study cohort as the standard population and age-specific standardised rates for population subgroups were calculated.[20] Sensitivity analysis was done using unstandardised rates.

This study adhered to the REporting of studies Conducted using Observational Routinely-collected Data reporting guidelines [21] (Supplementary Table 3). All data management, statistical analyses, and plotting was done in R version 3.6.2 [22], available within in the ISO27001 and Scottish Government approved Health Informatics Centre Safe Haven. The analysis was approved by CPRD Independent Scientific Advisory Committee (reference 20_018) under the terms of CPRD NHS Research Ethics dataset approval.

## Results

The study included 1168620 people. When considering all 80 conditions in the count, multimorbidity was present in 473533 (40.5%) of the cohort. People with multimorbidity were older than the whole population, median 60 years (IQR 46-72) versus 44 years (IQR 23-60), more often women, 257237 (54.3%) versus 587687 (50.3%), and more often lived in the five most deprived IMD decile areas, 208386 (44.0%) versus 505322 (43.2%). Differences between the whole population and people with multimorbidity examined using χ^2^ tests for proportions within each age-group, IMD decile, and both sexes, were statistically significant (p <0.001) (Table 1).

**Table 1.** Population characteristics.

|  | **No. (% of total population/each column)**<br>**N=1168620** | **No. (% of each row) with multimorbidity when considering all 80 conditions in the count** |
| --- | --- | --- |
| **Age group, years\*** |  |  |
| 0 – 9 | 113955 (9.8) | 2739 (2.4) |
| 10 – 19 | 137517 (11.8) | 9129 (6.6) |
| 20 – 29 | 122237 (10.5) | 24916 (20.4) |
| 30 – 39 | 143243 (12.3) | 39888 (27.8) |
| 40 – 49 | 176061 (15.1) | 66878 (37.9) |
| 50 – 59 | 173435 (14.8) | 89586 (51.5) |
| 60 – 69 | 141041 (12.1) | 98512 (69.9) |
| 70 – 79 | 97843 (8.4) | 82752 (84.4) |
| ≥80 | 63288 (5.4) | 59092 (93.4) |
| **Sex\*** |  |  |
| Men | 580933 (49.7) | 215555 (37.1) |
| Women | 587687 (50.3) | 257049 (43.7) |
| **IMD decile\*, \*\*** |  |  |
| 1 (most affluent) | 167558 (14.0) | 62032 (37.0) |
| 2 | 129704 (11.0) | 51504 (39.7) |
| 3 | 128234 (11.0) | 51794 (40.4) |
| 4 | 109986 (9.4) | 45681 (41.5) |
| 5 | 127816 (11.0) | 53601 (41.9) |
| 6 | 104158 (8.9) | 44279 (42.5) |
| 7 | 108782 (9.3) | 44097 (40.5) |
| 8 | 103501 (8.9) | 43102 (41.6) |
| 9 | 100577 (8.6) | 40019 (39.8) |
| 10 (most deprived) | 88304 (7.6) | 36495 (41.3) |

\*There were statistically significant differences in the proportion of people with and without multimorbidity ( $P < 0.001$ ) within each variable (row) group using $\chi^2$ tests.
\*\*Index of multiple deprivation (IMD)

Of the 80 conditions examined, six conditions were present in more than 5% of the whole population: hypertension was the most prevalent (9.1%), followed by depression (8.7%), asthma (7.6%), upper gastro-intestinal (GI) tract acid conditions (7.6%), anxiety (6.7%), and osteoarthritis (5.7%). Nine conditions were present in less than 0.1% of the population (Table 2). There was marked variability in multimorbidity prevalence depending on the number of conditions considered in the count. Using all 80 conditions, multimorbidity prevalence was 40.5% (95%CI [40.4,40.6] p <0.001). When considering only the two conditions most prevalent in the whole population in the count, multimorbidity was present in 4.6 % (95%CI [4.6,4.6] p <0.001) (Figure 1, Supplementary Table 3). When adding more conditions to the count (the most prevalent remaining conditions first), there was a steep increase in estimated multimorbidity prevalence, rising to 29.5% (95%CI [29.5,29.6%] p <0.001) when considering ten conditions in the count. Following this, a more gradual increase in estimated prevalence was seen as more conditions were added to the count: 35.2 % (95%CI [35.1,35.3] p <0.001) considering 20 conditions, and 37.4% (95%CI [37.3,37.5] p <0.001) considering 30 conditions. There was only 0.7 percentage point absolute difference in prevalence between considering 50 conditions, 39.8% (95%CI [39.7,39.9] p <0.001), and all 80 conditions (40.5%). In the whole population, the predefined ceiling where adding additional conditions had little impact on prevalence was reached at 52 conditions (i.e., estimated prevalence for 52 conditions versus 80 conditions relative risk [RR] >0.99).

**Figure 1.**
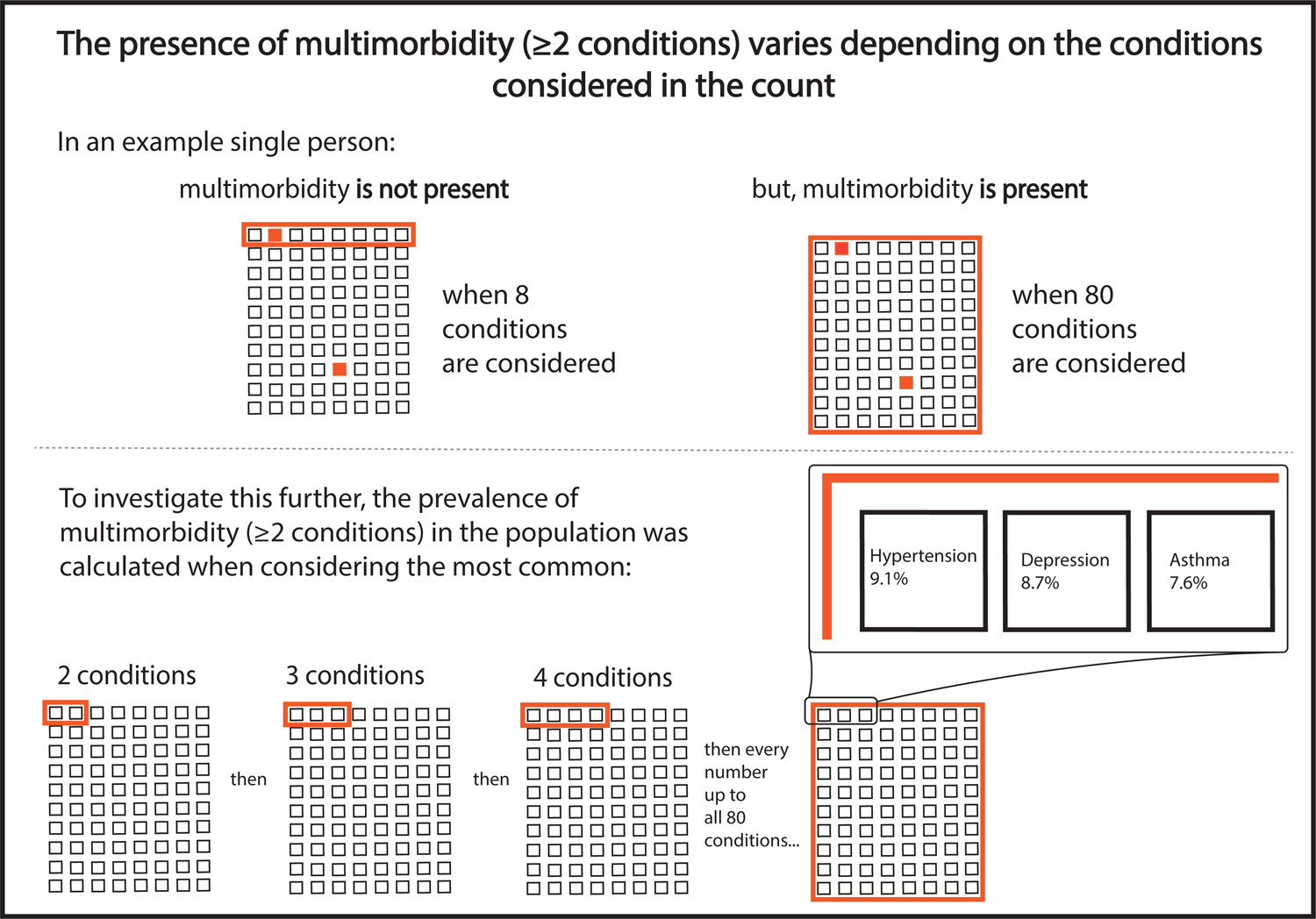
Multimorbidity prevalence according to number of conditions, the ceiling effect where adding additional conditions had little impact on prevalence, and selection of conditions using existing condition-lists. The black line represents multimorbidity prevalence calculated when considering different numbers of conditions in the count ranging from two to all 80 conditions, where conditions were added in order of most to least prevalent (e.g., at two conditions this is multimorbidity prevalence considering the most common two conditions). Percentage prevalence of multimorbidity when 10, 20, 30, 40, 50, 60, 70, and 80 conditions were considered marked at empty black circles above the black line. The number of conditions at which relative risk (RR) was >0.99 of multimorbidity prevalence of having the same multimorbidity prevalence when all 80 conditions were considered (ceiling effect) was reached is marked with an orange dot (at 52 conditions). Black dots represent multimorbidity prevalence when considering conditions included in existing condition-lists and are annotated with the condition-list name, prevalence, and number of conditions considered.

**Table 2.** Prevalence of individual conditions.

| Prevalence rank | Long-term condition | Population count (%)<br>N=1168620 | Prevalence rank | Long-term condition | Population count (%)<br>N=1168620 |
| --- | --- | --- | --- | --- | --- |
| 1 | Hypertension | 212520 (18.2) | 41 | Paroxysmal tachycardias | 8747 (0.7) |
| 2 | Depression | 201991 (17.3) | 42 | Obstructive and reflux uropathy | 8501 (0.7) |
| 3 | Asthma | 177301 (15.2) | 43 | Polymyalgia rheumatica | 8446 (0.7) |
| 4 | Upper gastro-intestinal tract acid conditions | 176518 (15.1) | 44 | Intellectual disability | 8004 (0.7) |
| 5 | Anxiety | 156762 (13.4) | 45 | Secondary malignancy | 7332 (0.6) |
| 6 | Osteoarthritis | 132799 (11.4) | 46 | Schizophrenia | 7097 (0.6) |
| 7 | Primary malignancy | 74917 (6.4) | 47 | Haematological malignancy | 7028 (0.6) |
| 8 | Type 2 diabetes mellitus | 61671 (5.3) | 48 | Autism | 6714 (0.6) |
| 9 | Chronic kidney disease | 59812 (5.1) | 49 | Visual impairment and blindness | 5942 (0.5) |
| 10 | Coronary heart disease | 58585 (5.0) | 50 | Obsessive compulsive disorder | 5553 (0.5) |
| 11 | Thyroid disease | 57289 (4.9) | 51 | Bronchiectasis | 5123 (0.4) |
| 12 | Erectile dysfunction | 53198 (4.5) | 52 | Bipolar affective disorder | 4572 (0.4) |
| 13 | Diverticular disease | 46410 (4.0) | 53 | Type 1 diabetes mellitus | 4439 (0.4) |
| 14 | Urinary incontinence | 45681 (3.9) | 54 | Coeliac disease | 4188 (0.4) |
| 15 | Psoriasis | 41871 (3.6) | 55 | Eating disorders | 3766 (0.3) |
| 16 | Benign prostatic hyperplasia | 33656 (2.7) | 56 | Tubulointerstitial nephropathy | 3511 (0.3) |
| 17 | Gout | 33483 (2.9) | 57 | Cardiomyopathy | 3399 (0.3) |
| 18 | Atrial fibrillation | 33098 (2.8) | 58 | Parkinson's disease | 2938 (0.2) |
| 19 | Alcohol misuse | 31841 (2.7) | 59 | Multiple sclerosis | 2932 (0.2) |
| 20 | Chronic obstructive pulmonary disease | 31654 (2.7) | 60 | Hyperparathyroidism | 2316 (0.2) |
| 21 | Osteoporosis | 30284 (2.6) | 61 | Ankylosing spondylosis | 2214 (0.2) |
| 22 | Stroke and transient ischaemic attack | 27925 (2.4) | 62 | Cerebral palsy | 2051 (0.2) |
| 23 | Urolithiasis | 25551 (2.2) | 63 | Lupus erythematosus | 1752 (0.1) |
| 24 | Peripheral neuropathy | 24065 (2.1) | 64 | Pulmonary fibrosis | 1712 (0.1) |
| 25 | Hearing loss | 22535 (1.9) | 65 | Primary thrombocytopaenia | 1707 (0.1) |
| 26 | Venous thromboembolic disease | 21058 (1.8) | 66 | Giant cell arteritis | 1662 (0.1) |
| 27 | Epilepsy | 19139 (1.6) | 67 | Paralysis | 1625 (0.1) |
| 28 | Heart valve disease | 18172 (1.5) | 68 | Primary pulmonary hypertension | 1210 (0.1) |
| 29 | Heart failure | 17879 (1.5) | 69 | Sjogren syndrome | 1184 (0.1) |
| 30 | Substance misuse | 17433 (1.5) | 70 | Sick sinus syndrome | 1097 (0.1) |
| 31 | Inflammatory arthritis | 15853 (1.4) | 71 | Thalassaemia | 1088 (0.1) |
| 32 | Endometriosis | 14567 (1.2) | 72 | Human immunodeficiency virus | 960 (0.1) |
| 33 | Sleep apnoea | 12853 (1.1) | 73 | Diabetes mellitus other or not specified | 933 (0.1) |
| 34 | Raynaud's disease | 12312 (1.0) | 74 | Aplastic anaemia | 820 (0.1) |
| 35 | Neuropathic bladder | 11507 (1.0) | 75 | Cystic fibrosis | 559 (0.05) |
| 36 | Dementia | 11374 (1.0) | 76 | Scleroderma | 442 (0.04) |
| 37 | Inflammatory bowel disease | 10949 (0.9) | 77 | Myasthenia gravis | 419 (0.04) |
| 38 | Liver disease | 10628 (0.9) | 78 | Sickle cell disease | 415 (0.04) |
| 39 | Peripheral arterial disease | 10427 (0.9) | 79 | Addison's disease | 394 (0.03) |
| 40 | Heart block & bundle branch block | 10231 (0.9) | 80 | Motor neurone disease | 231 (0.02) |

**Table 3.** Multimorbidity prevalence using existing condition-lists and relative risk (RR) of multimorbidity when considering all 80 conditions (ceiling effect where adding additional conditions had little impact on prevalence).

| Condition-list | No. of conditions in condition-list | Multimorbidity prevalence<br>No. (%) | RR (95% CI) |
| --- | --- | --- | --- |
| All 80 conditions (reference) | 80 | 473533 (40.5) | 1.0 |
| Ho always + usually[10] | 55 | 425413 (36.4) | 0.90 (0.89-0.90) |
| Barnett[16] | 41 | 397009 (34.0) | 0.84 (0.83-0.84) |
| N'Goran[11] | 29 | 219098 (18.7) | 0.46 (0.46-0.46) |
| Elixhauser[19] | 28 | 268261 (23.0) | 0.57 (0.56-0.57) |
| Fortin[9] | 27 | 389286 (33.3) | 0.82 (0.82-0.82) |
| Ho always[10] | 27 | 129698 (11.1) | 0.27 (0.27-0.27) |
| Charlson[18] | 21 | 135166 (11.6) | 0.28 (0.28-0.29) |
| Diederichs[6] | 18 | 224001 (19.2) | 0.47 (0.47-0.47) |
| Salive[17] | 14 | 222859 (19.1) | 0.47 (0.47-0.47) |

Multimorbidity prevalence varied widely between the nine different condition-lists, varying from 11.1% (95%CI [11.0,11.2] p <0.001) using the Ho always[10] list, to 36.4% (95%CI [36.3,36.5] p <0.001) using the Ho always + usually [10] list (Figure 1, Table 2). Three condition-lists (Ho always + usually [10], Barnett [16], Fortin [9]) had prevalence close to that estimated by including the same number of the most common conditions in the number of conditions analysis (represented by proximity of these points to the black line in Figure 1). These lists also had highest RR of the multimorbidity prevalence calculated when considering all 80 conditions in the count: Ho always + usually [10] RR 0.90 (95%CI [0.89,0.90] p <0.001), Barnett et al [16] RR 0.84 (95%CI [0.83,0.84] p <0.001), and Fortin et al [9] RR 0.82 (95%CI [0.82,0.82] p <0.001) (Table 2). The remaining five condition-lists, however, had prevalence considerably below that estimated by including the same number of most common conditions with prevalence RR 0.27 (95%CI [0.27,0.27] p <0.001) and RR 0.27 (95%CI [0.27,0.27] p <0.001) respectively using the Ho always [10] and Charlson [18] condition-lists.

The initial gradient of increase in multimorbidity prevalence seen as conditions were added to the count was steepest in the oldest age-groups, followed by flattening of the curve as more conditions were considered (Figure 2). In 0-9- and 10–19-year-olds, there was a more gradual increase in prevalence, because rarer conditions contribute to a higher proportion of multimorbidity in children and young people. The influence of adding additional numbers of conditions to the count on estimated prevalence plateaued at a lower number of conditions considered in older people. In people aged 80 years and over, the predefined ceiling (prevalence compared to 80 conditions RR >0.99) was reached at 29 conditions, compared to 71 conditions in those aged 0-9 years (Figure 2). In IMD-stratified analysis, there was a clear social gradient of multimorbidity prevalence irrespective of the number of conditions included, with the more deprived having higher prevalence than the less deprived (Figure 3). The predefined ceiling was reached at a lower number of conditions in the most deprived IMD decile (49 conditions) compared to the least deprived (54 conditions) (Figure 3). Sensitivity analysis using raw (unstandardised rates) had less clear social gradient (reflecting that the most deprived are on average younger than the affluent), and no clear pattern in the predefined ceiling across IMD deciles (Supplementary Figure 1).

**Figure 2.**
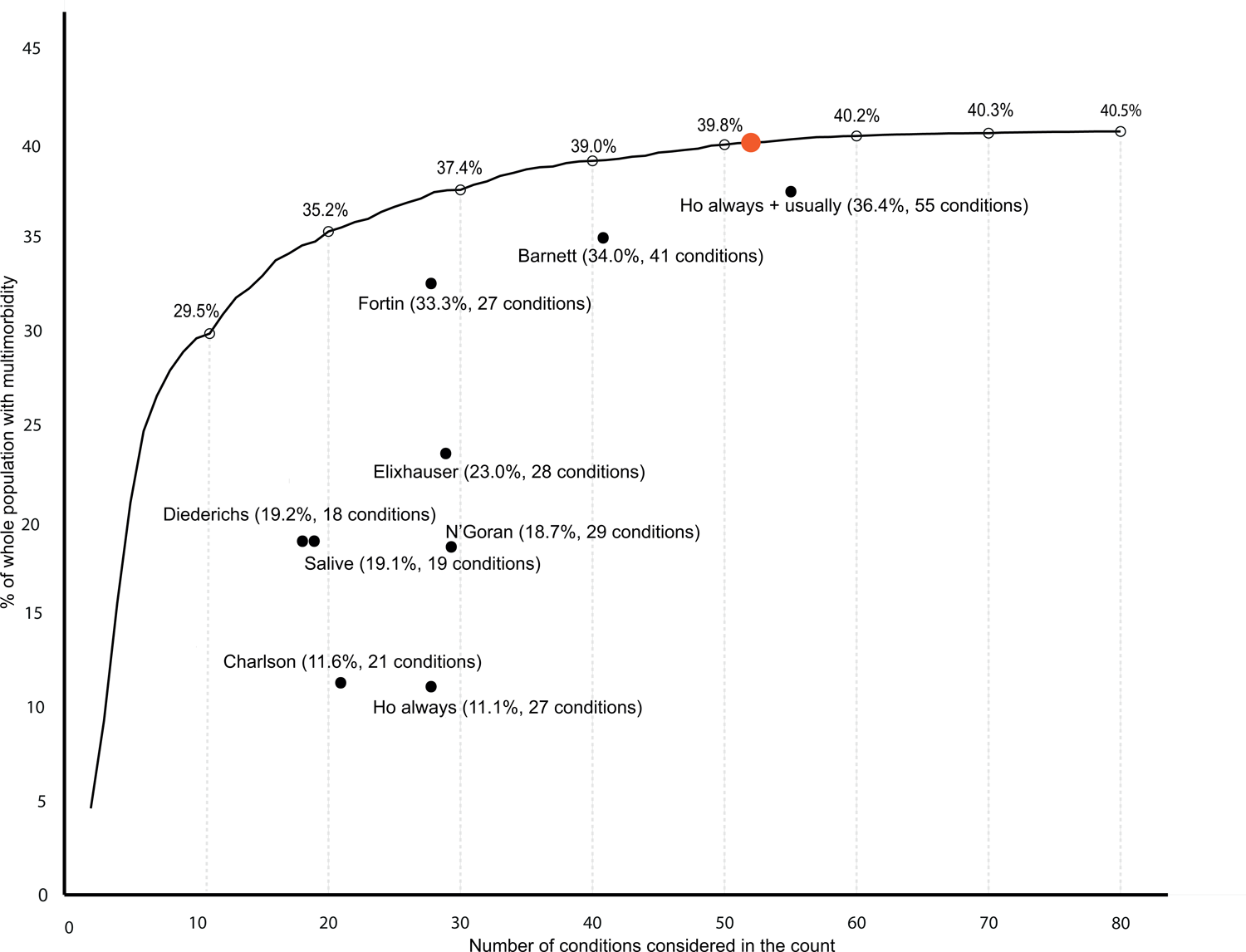
Age-stratified multimorbidity prevalence according to number of conditions considered, reporting the ceiling effect where adding additional conditions had little impact on prevalence. Labelled coloured lines represent multimorbidity prevalence calculated when considering different numbers of conditions in the count ranging from two to all 80 conditions stratified into age groups. Black dots represent the number of conditions at which relative risk (RR) >0.99 of multimorbidity prevalence of having the same multimorbidity prevalence when all 80 conditions were considered (ceiling effect): 0-9 years at 71 conditions, 10-19 years at 67 conditions, 20-29 conditions at 57 conditions, 30-39 years at 57 conditions, 40-49 years at 56 conditions, 50-59 years at 50 conditions, 60-69 years at 44 conditions, 70-79 years at 37 conditions, 80+ years at 29 conditions.

**Figure 3.**
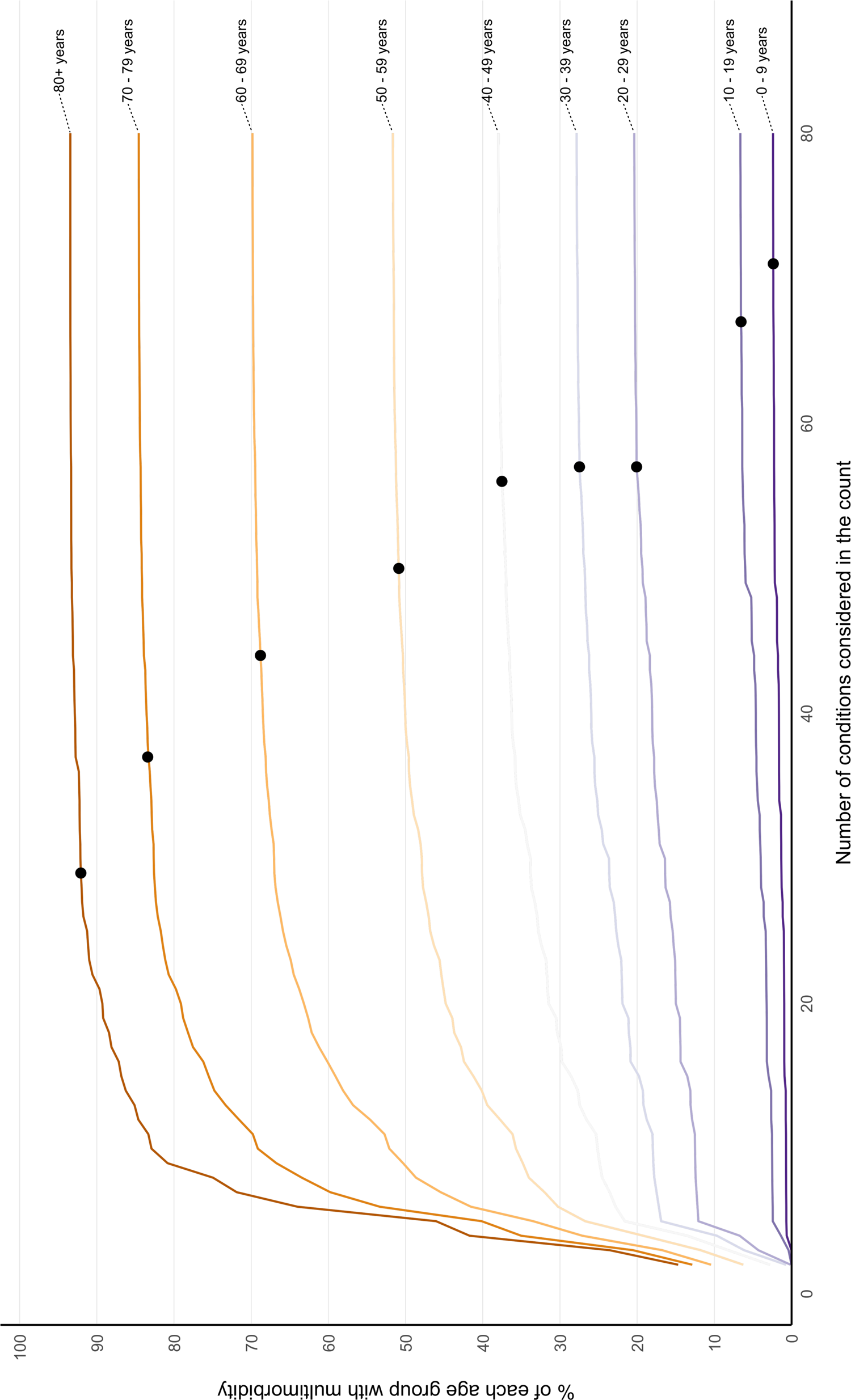
Socioeconomic position stratified multimorbidity prevalence according to number of conditions considered following direct age standardisation, reporting the ceiling effect where adding additional conditions had little impact on prevalence. Labelled coloured lines represent multimorbidity prevalence calculated when considering different numbers of conditions in the count ranging from two to all 80 conditions stratified into Index of Multiple Deprivation (IMD) deciles where IMD 1 is least and IMD 10 is most deprived. Black dots represent the number of conditions at which relative risk (RR) >0.99 of multimorbidity prevalence of having the same multimorbidity prevalence when all 80 conditions were considered (ceiling effect): IMD 10 at 49 conditions, IMD 9 at 50 conditions, IMD 8 at 50 conditions, IMD 7 at 51 conditions, IMD 6 at 51 conditions, IMD 5 at 53 conditions, IMD 4 at 52 conditions, IMD 3 at 53 conditions, IMD 2 at 53 conditions, and IMD 1 at 54 conditions. Direct age standardisation where the whole study cohort was the standard population was applied (see Supplementary Figure 1 for unstandardised rates).

Multimorbidity prevalence was higher in women and girls at every level of number of conditions in the direct age-standardised analysis, and the predefined ceiling was reached at a higher number of conditions in women and girls than in men and boys (Figure 4). In the sensitivity analysis using unstandardised rates, there was a larger gap in multimorbidity prevalence between sexes, reflecting that women are on average older, and the predefined ceiling was reached at a similar number of conditions to the direct age-standardised analysis (Supplementary Figure 2).

**Figure 4.**
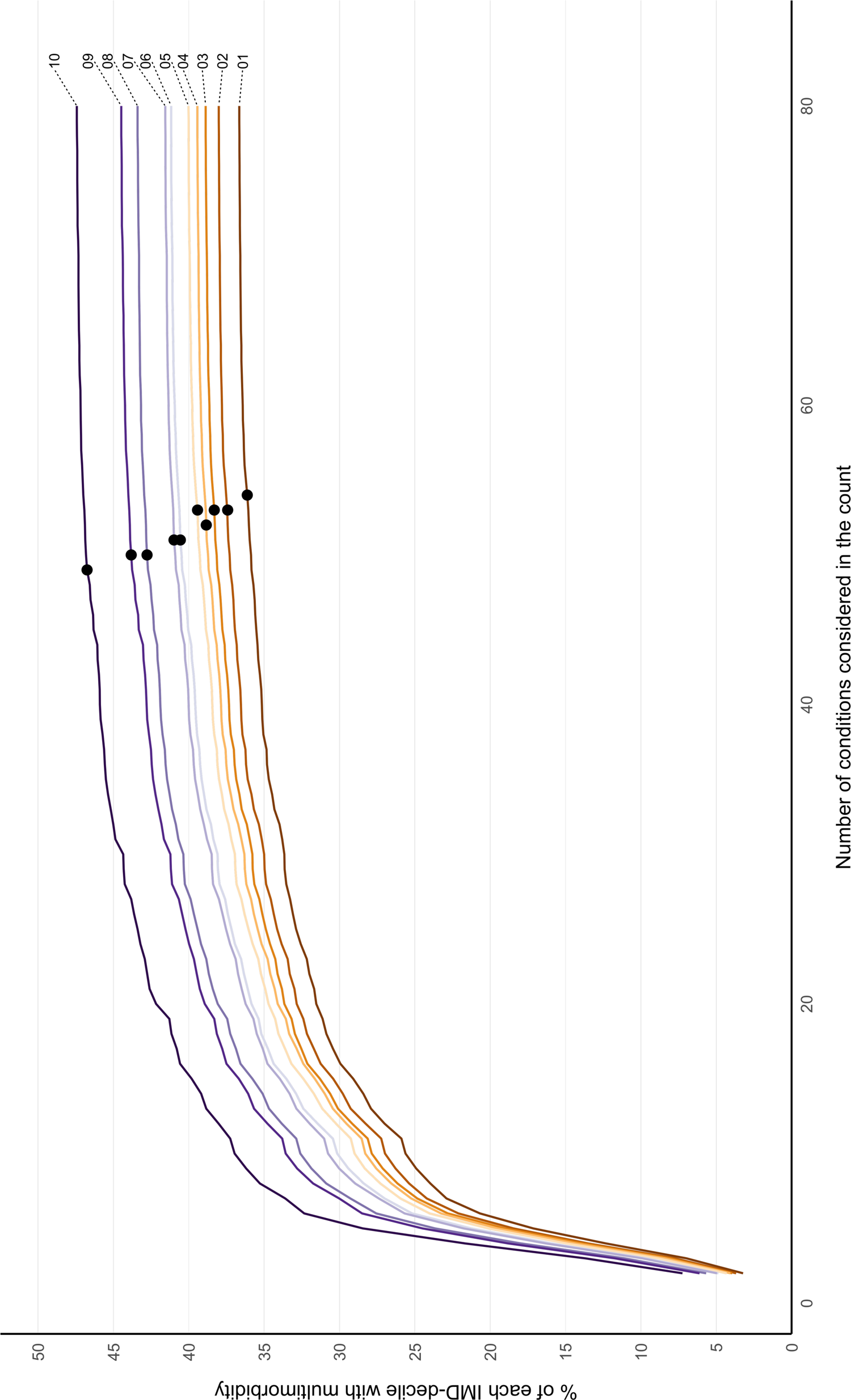
Sex stratified multimorbidity prevalence according to number of conditions considered following direct age standardisation, reporting the ceiling effect where adding additional conditions had little impact on prevalence. Labelled coloured lines represent multimorbidity prevalence calculated when considering different numbers of conditions in the count ranging from two to all 80 conditions stratified by sex. Black dots represent the number of conditions at which relative risk (RR) >0.99 of multimorbidity prevalence of having the same multimorbidity prevalence when all 80 conditions were considered (ceiling effect): women and girls at 54 conditions and men and boys at 50 conditions. Direct age standardisation where the whole study cohort was the standard population was applied (see Supplementary Figure 2 for unstandardised rates).

**Figure 5.**
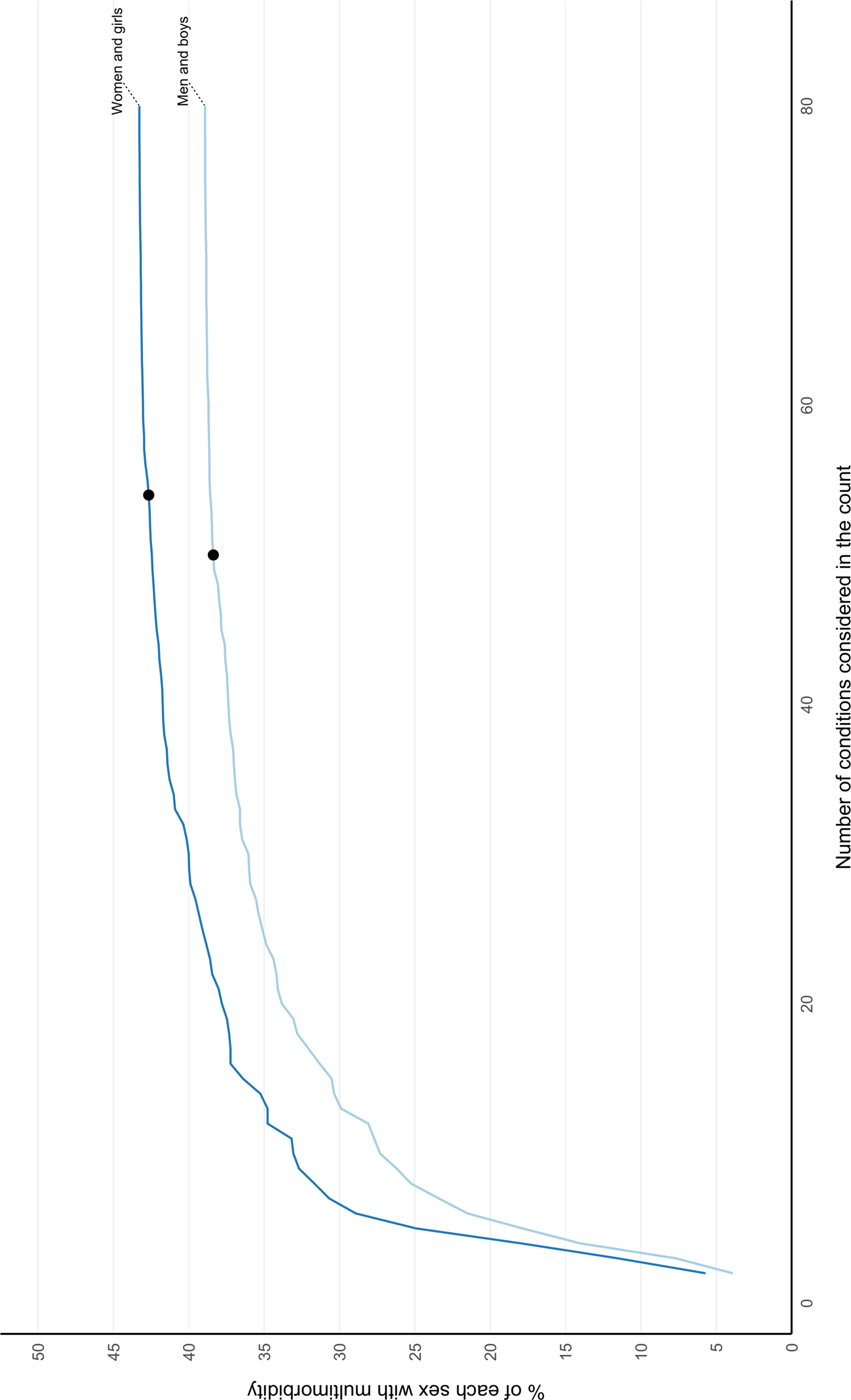
Multimorbidity prevalence by age considering all 80 conditions and according to existing condition-lists. Labelled coloured lines represent multimorbidity prevalence calculated for each age group when considering conditions in each condition-list.

In both age and deprivation stratified analyses, fewer conditions were required to reach RR 0.99 for the groups with highest prevalence. However, a different pattern was seen in men (who had lower multimorbidity prevalence) where the ceiling was reached at 50 conditions, compared to 54 in women (Figure 4).

The age-distribution of multimorbidity prevalence was not uniform across the nine condition-lists. Across all ages, multimorbidity prevalence using the Ho always + usually [10] condition-list was closest to prevalence when considering all 80 conditions (Figure 4). The Fortin [9], Barnett [16], and Elixhauser [19] condition-lists had lower prevalence than Ho always + usually [10] but followed a similar upward trajectory from youngest to oldest. Salive et al [17] and Diederichs et al [6] had low prevalence in younger age-groups, but multimorbidity prevalence increased steeply from age 50-59 years and older onwards. Ho always [10] and Charlson [18] had markedly lower prevalence rates than other condition-lists across all age-groups.

## Discussion

This study found very large differences in estimated multimorbidity prevalence from varying the number and selection of conditions considered in the count, and in younger people, the more affluent, and women, including additional relatively rare conditions had larger impact on estimated multimorbidity prevalence. Multimorbidity prevalence differed considerably by varying the number of conditions, ranging from 4.6% to 40.5%, and selection of conditions considered, ranging from 11.1% to 36.4% using nine previously published lists of conditions.[6, 9, 10, 16–19] Counting multimorbidity prevalence using the nine existing condition-lists resulted in lower estimated prevalence than when considering the same number of the most common conditions, although the extent of this varied: Ho always + usually [10], Fortin [9], and Barnett [16] had the best performance.

Consistent with the findings of this study, there is a wide range in the number and selection of conditions considered in the current multimorbidity literature [4], and in estimates of multimorbidity prevalence [7]. A systematic review of 566 multimorbidity studies by Ho et al [4] found that the number of conditions considered by existing studies ranged from two to 285 (median 17, IQR 11-23), and very little uniformity in terms of the selection of conditions was found across studies. Only eight conditions (diabetes, stroke, cancer, chronic obstructive pulmonary disease, hypertension, coronary heart disease, chronic kidney disease, and heart failure) were considered in at least half of the studies, and a quarter of studies did not consider any mental health condition. Simard et al [23] reviewed existing literature to examine how studies used, developed, and validated methods for measuring multimorbidity. They found heterogeneity in the grouping of conditions, validation processes, number of ICD-10 code digits used to define included conditions, and use of additional data sources. Diederichs et al [6] recognised the need to establish a standardised instrument to measure multimorbidity, and recommended a minimum set of 11 conditions to include (cancer, diabetes mellitus, depression, hypertension, myocardial infarction, chronic ischemic heart disease, heart arrhythmias, heart insufficiency, stroke, COPD, and arthritis). These conditions were selected based on high prevalence and a severe impact on affected individuals in terms of impairment of function and high need for management, from a population of people aged over 64 years old in Germany. A recent systematic review and meta-analysis of 193 studies examining multimorbidity prevalence [7] did not directly compare prevalence when considering different condition-lists, however did find that prevalence was significantly higher in studies considering a larger number of conditions in the count: studies considering 44 or more conditions had higher pooled multimorbidity prevalence (87.6%) than studies considering fewer than nine conditions (30.1%).

Strengths of this study include comprehensive analysis of multimorbidity prevalence estimates in a large population dataset derived from primary care electronic health records. Analysis systematically examined multimorbidity prevalence in the same population for different numbers of conditions considered in the count and using condition-lists recommended or used in previous studies. However, a limitation is that we did not necessarily replicate how previous studies ascertained the presence of conditions, but instead defined the presence of each condition using published UK code-lists. This improves comparability within this study but highlights that further variability in prevalence estimates will happen because of variation in how each condition is ascertained (i.e., variation in exactly which codes or prescriptions are used, or restrictions on how recent a diagnosis must be). There is heterogeneity in which conditions are included between existing lists of conditions, and therefore decisions were made about how to standardise conditions to the full list of 80 conditions. For example, the Barnett (2012) [16] condition-list used time-limited diagnoses which were not replicated in this study in order to make condition ascertainment consistent across the condition-lists examined. Using electronic health records to ascertain the prevalence of conditions can be associated with under ascertainment because the absence of a record does not necessarily mean absence of the condition, and more severe disease is likely to be over-represented in medical records [24]. The data were from 2015 however prevalence of the commonest condition hypertension was unchanged between 1990 and 2019 in a pooled analysis of worldwide population studies (32% of women in 2019 versus 32% in 1990) [25], and this study identified similar prevalence rates of depression as ascertained by an Office of National Statistics survey from 2021 [26].

Deciding which conditions to include in multimorbidity research is complicated, including in the extent to which conditions should be aggregated (e.g., coronary heart disease) or considered separately (e.g., angina, myocardial infarction). Ideally, researchers would use a standardised list to improve research comparability and reproducibility, but this isn’t always feasible due to varying data availability and varying prevalence of disease in different settings. An alternative method is to use an “open condition-list”, as used by Fortin et al [8] in a Canadian study where methodology was not constrained to a specified number of conditions considered in the count to calculate multimorbidity prevalence, but considered all conditions present in a patient’s medical records in the multimorbidity count. The number of conditions in study participants was highly variable and resulted in large differences in multimorbidity prevalence, particularly for younger people [7]. The method of data collection involved manual review of medical records, and therefore although this analysis provides additional richness, it would not be easy to scale this approach and apply it to larger populations.

Even where researchers agree on which conditions to measure, there is an additional source of variation introduced by heterogeneity in methods chosen to measure and ascertain those conditions in data. Based on this research, if the purpose of the study is to estimate prevalence then estimates will be relatively stable providing the 50 most common conditions are considered, although this threshold requires examination in other datasets and settings. Although some tailoring to local context and purpose will often be necessary, comparability and reproducibility would be improved by choices always starting with a core list of conditions. Researchers should therefore consider using the Delphi consensus derived Ho always + usually list [10], or for measuring prevalence the Barnett [16], or Fortin [9] condition-lists.

There are several areas where further research is needed. First, this study examined relationships between the number and selection of conditions and multimorbidity prevalence in the UK. However, similar studies in low- and middle-income countries are needed, where prevalence of individual conditions will be different. Second, condition ascertainment in routine data is based on lists of clinical codes (and sometimes prescribing or laboratory data) [27]. However, there can be large variations in the clinical codes used to define the same condition in different studies [28]. Therefore, further exploration of the impact of variation in which codes or prescriptions are used to define conditions is needed. Applying the condition codes from validated open-access published code lists, such as the HDR-UK Phenotype Library [15], or other similar sources [29] will also improve comparability and reproducibility. Third, it is important to examine whether and how much the number and selection of conditions considered in counts alter observed associations with important clinical outcomes such as functional status, unplanned hospital use, and death.

The key implication of this study is that the choice of conditions to consider when estimating multimorbidity prevalence has a large impact on the results, with additional variation in impact between older versus younger people particularly. The comparability and reproducibility of multimorbidity research would be improved by researchers including recommended core conditions wherever possible [10], with explicitly justified variation for study context and purpose.

## Author contributions

CM and BG conceptualised the study, contributed to the design of the project, and wrote the first draft. CM analysed the data. CM, SWM, and BG interpreted the results. MM and EJ cleaned the dataset, and MM and DH provided support with the analysis. BG, SWM, DM, EJ, DM acquired the funding for the dataset. CM, MM, DH, DAM, EJ, CD, JL, RAL, IH, SWM, and BG contributed to interpretation of the data, and critically commented on the manuscript. CM, MM, DH, DAM, EJ, CD, JL, RAL, IH, SWM, and BG reviewed drafts and approved the final version.

## Data Availability

All relevant data are within the manuscript or available via GitHub repository at https://github.com/macraec. Raw data cannot be made publicly available to protect data security.

## Information Panel

### Supplementary File

Measuring multimorbidity: impact of varying the number and selection of conditions on estimated multimorbidity prevalence in a large primary care population dataset

## Supplementary Information Panel. Measuring multimorbidity in research: a Delphi consensus study

Two recommended condition-lists defined by a modified Delphi panel study[10] were used. This study developed international consensus on the measurement of multimorbidity in research and was funded by Health Data Research UK (HDR-UK). Data were collected in three rounds of online questionnaires, including 25 public panel and 150 professional panel members. Public members had an interest in, or personal experience of, multimorbidity. Professional participants were clinicians, policy makers, and researchers interested in, or involved in, multimorbidity work. Two sets of questions were developed separately for the public and professional panels. Round one questions were informed by a recent systematic review.[30] Questions in subsequent rounds were informed by results from the previous questionnaire. Participants were asked to answer open and closed questions, where open questions were subsequently triangulated by subsequent closed questions. Consensus was reached for 24 conditions to always include in multimorbidity measures, and 35 conditions to usually include unless a good reason not to, and these lists have been examined in our study of multimorbidity prevalence. In the study, we calculated multimorbidity prevalence using the always include list (Ho always), and both condition-lists together (Ho always + usually).

---

**Supplementary Figure 1.**
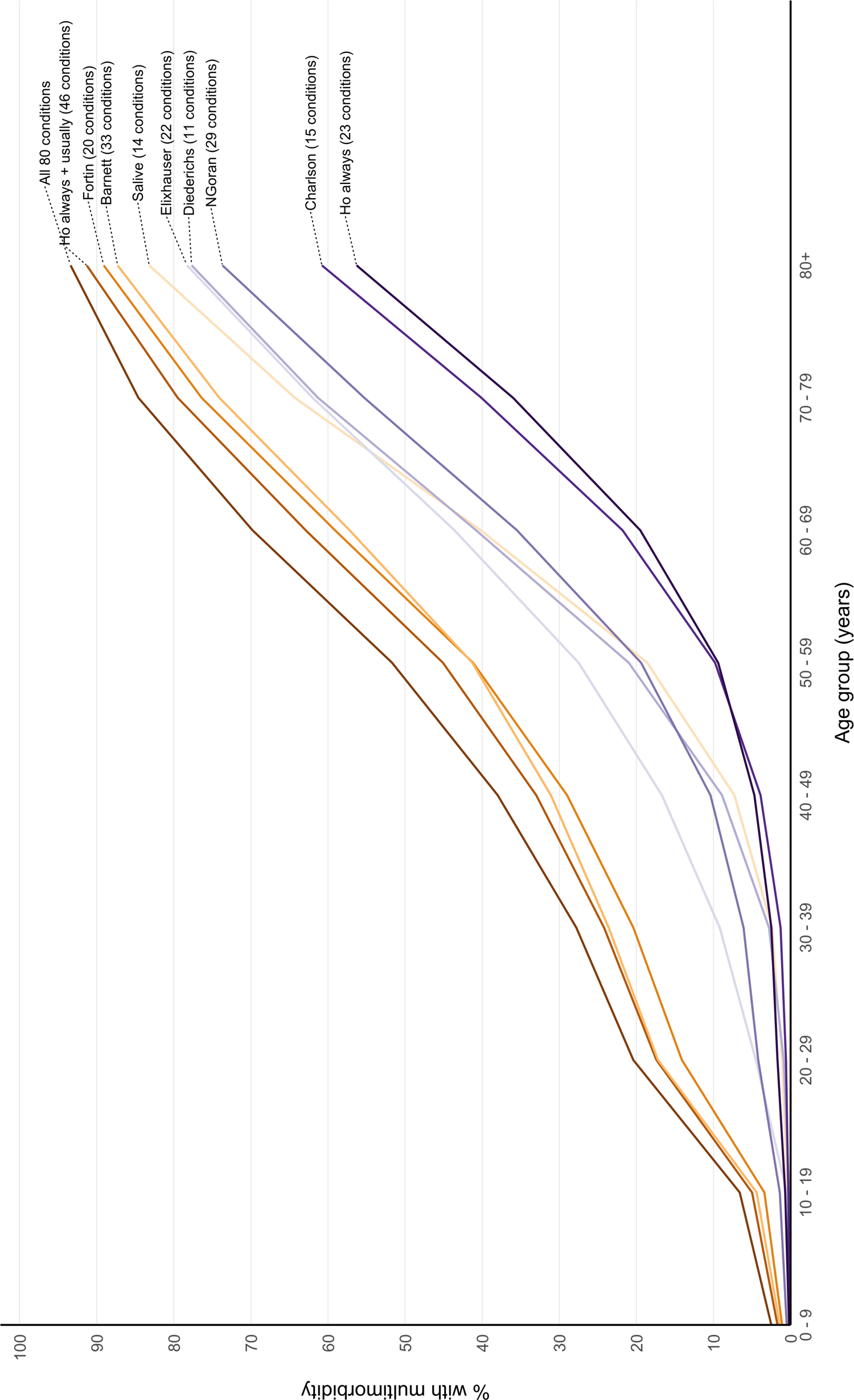
Socioeconomic position stratified multimorbidity prevalence according to number of conditions considered without direct age standardisation, reporting the ceiling effect where adding additional conditions had little impact on prevalence. Labelled coloured lines represent multimorbidity prevalence calculated when considering different numbers of conditions in the count ranging from two to all 80 conditions stratified into Index of Multiple Deprivation (IMD) deciles where IMD 1 is least and IMD 10 is most deprived. Black dots represent the number of conditions at which relative risk (RR) >0.99 of multimorbidity prevalence of having the same multimorbidity prevalence when all 80 conditions were considered (ceiling effect): IMD 10 at 51 conditions, IMD 9 at 53 conditions, IMD 8 at 51 conditions, IMD 7 at 51 conditions, IMD 6 at 51 conditions, IMD 5 at 52 conditions, IMD 4 at 51 conditions, IMD 3 at 52 conditions, IMD 2 at 52 conditions, and IMD 1 at 54 conditions.

**Supplementary Figure 2.**
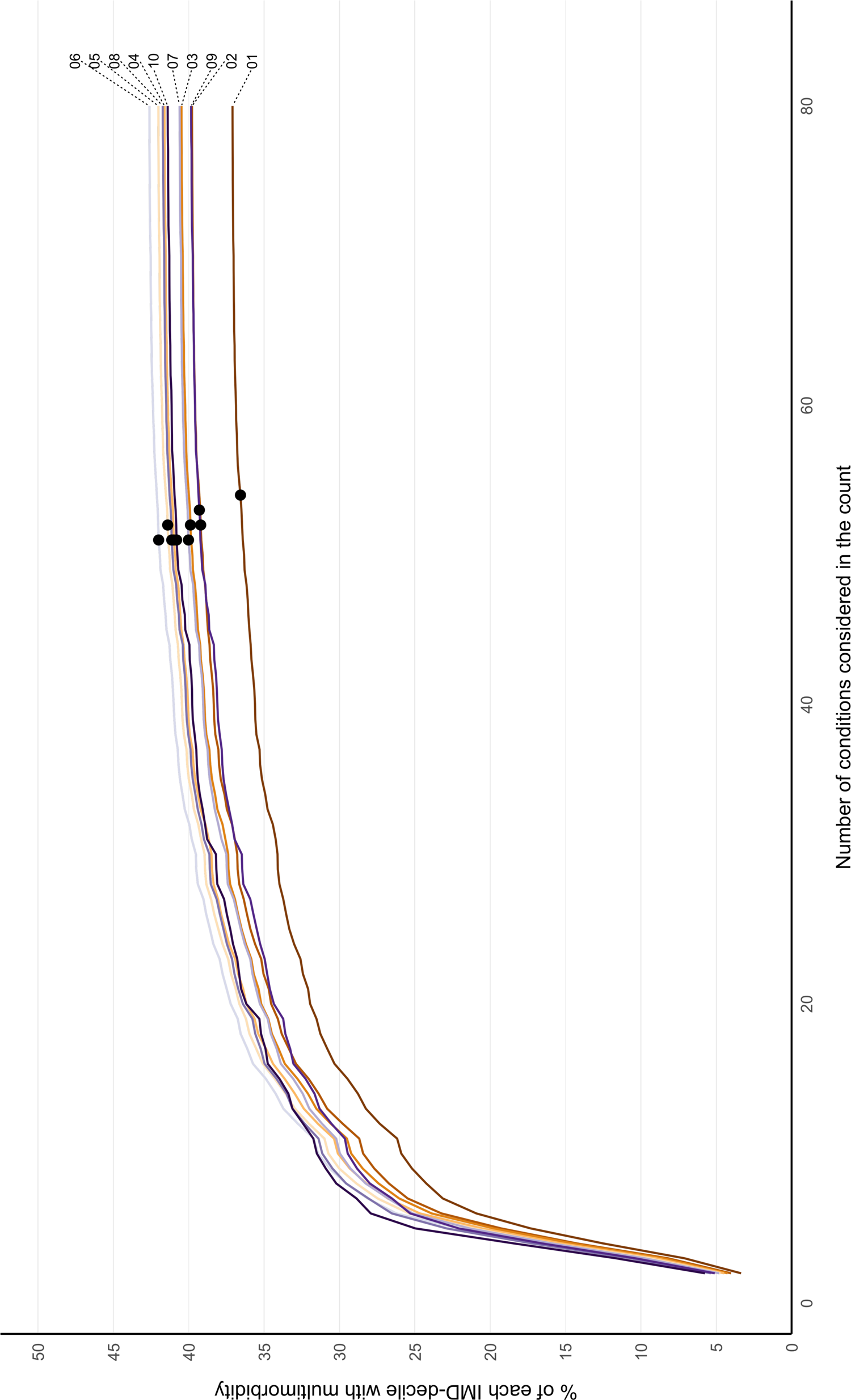
Sex stratified multimorbidity prevalence according to number of conditions considered without direct age standardisation, reporting the ceiling effect where adding additional conditions had little impact on prevalence. Labelled coloured lines represent multimorbidity prevalence calculated when considering different numbers of conditions in the count ranging from two to all 80 conditions stratified by sex. Black dots represent the number of conditions at which relative risk (RR) >0.99 of multimorbidity prevalence of having the same multimorbidity prevalence when all 80 conditions were considered (ceiling effect): women and girls at 54 conditions and men and boys at 51 conditions.

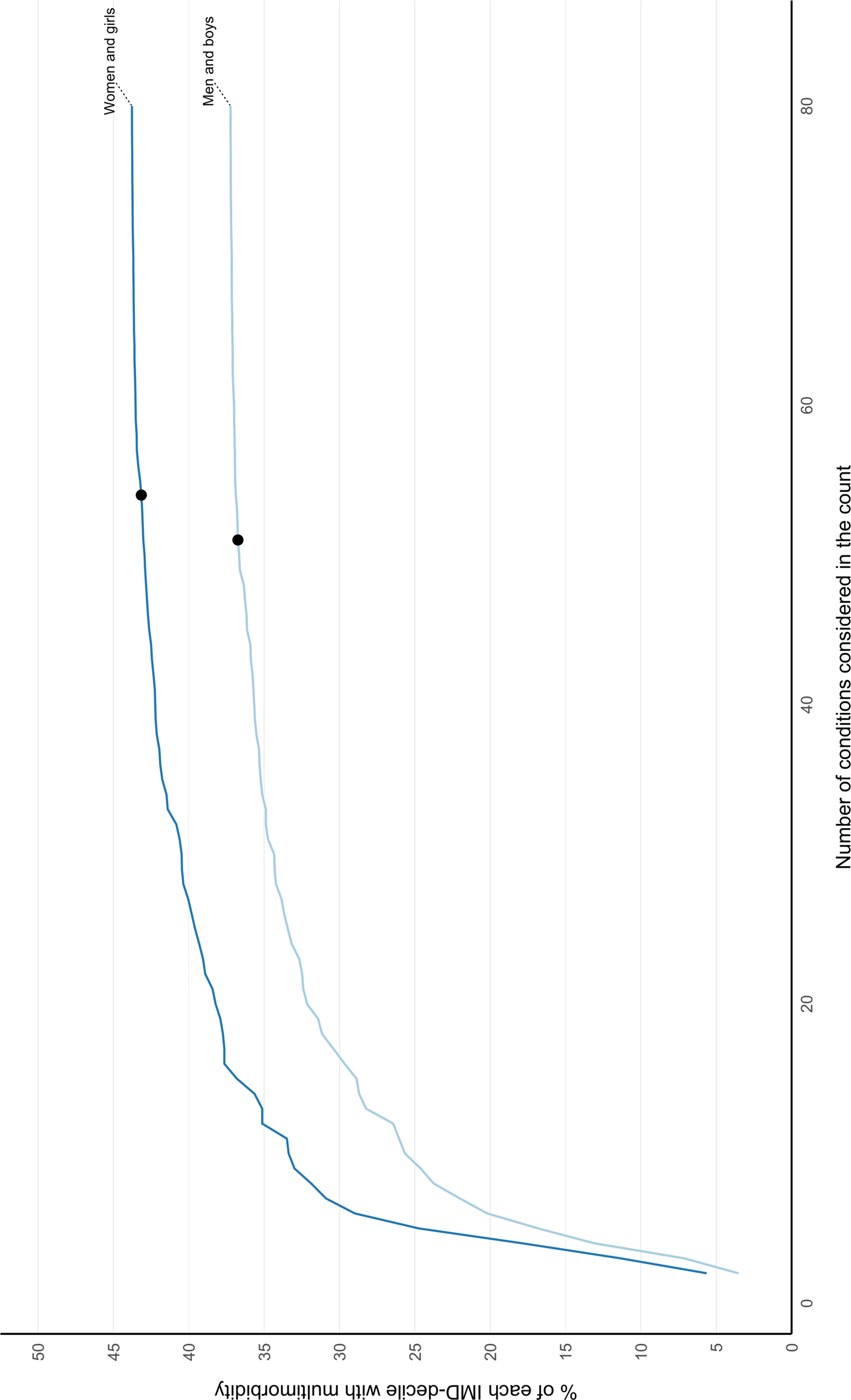

**Supplementary Table 1.**
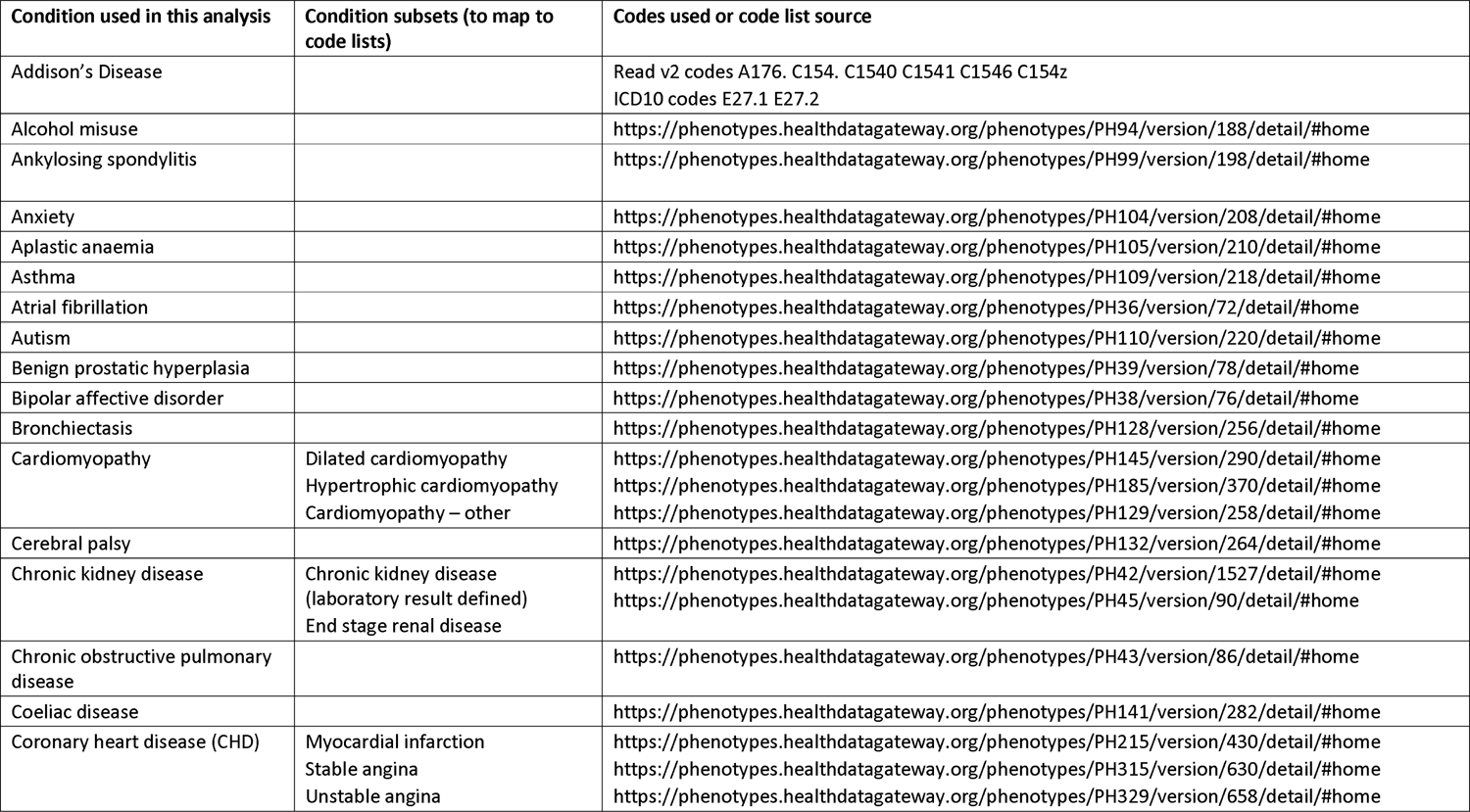
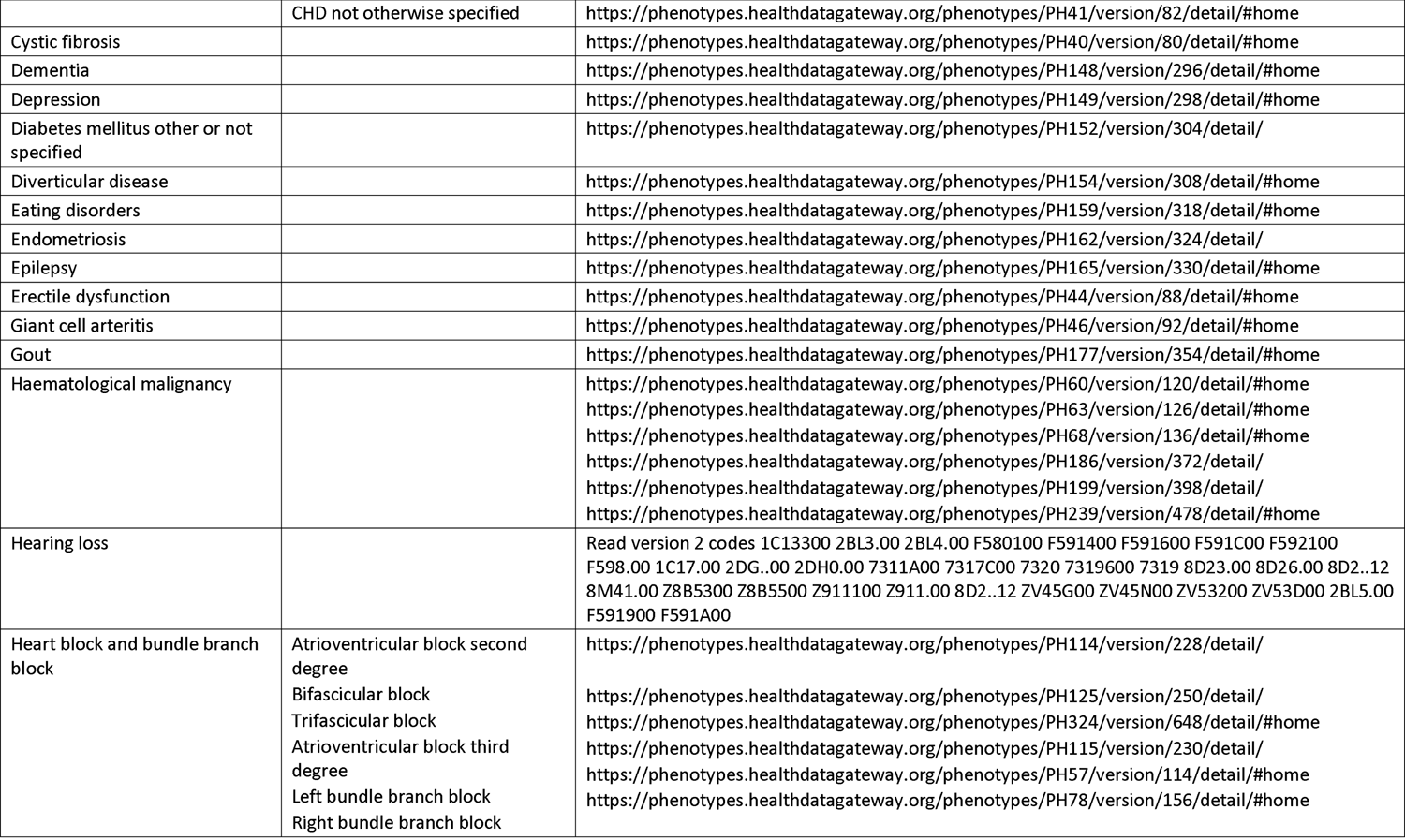
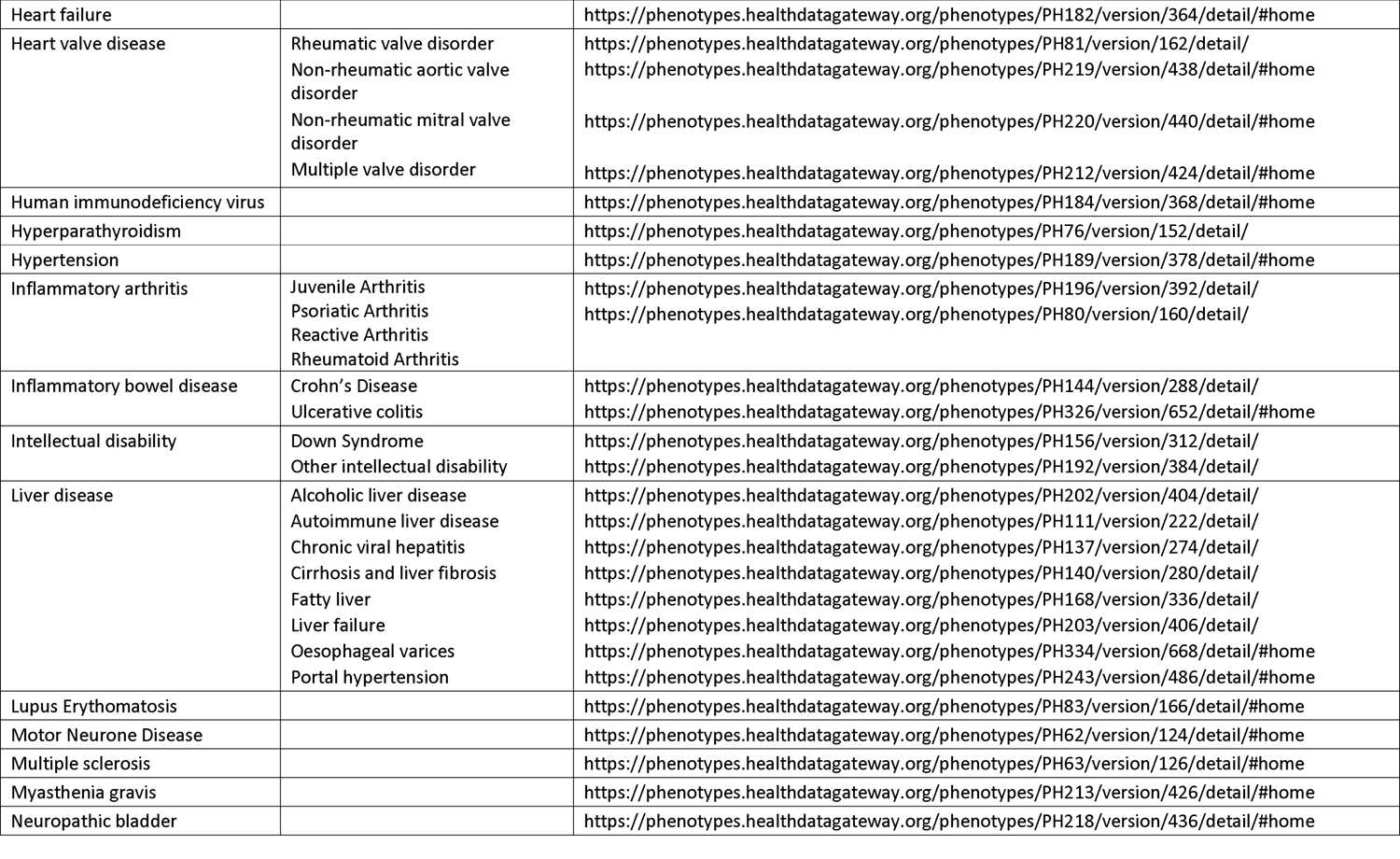
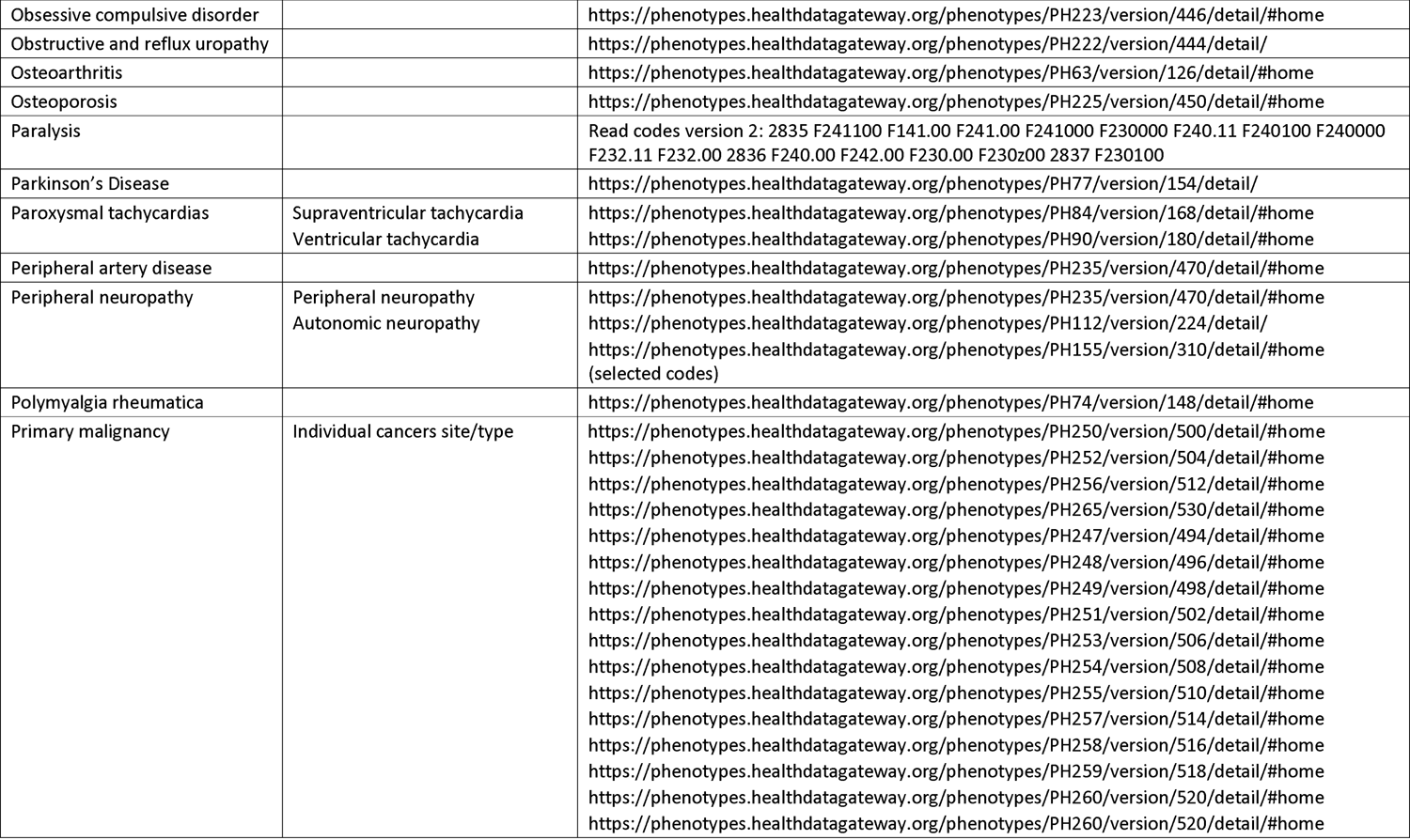
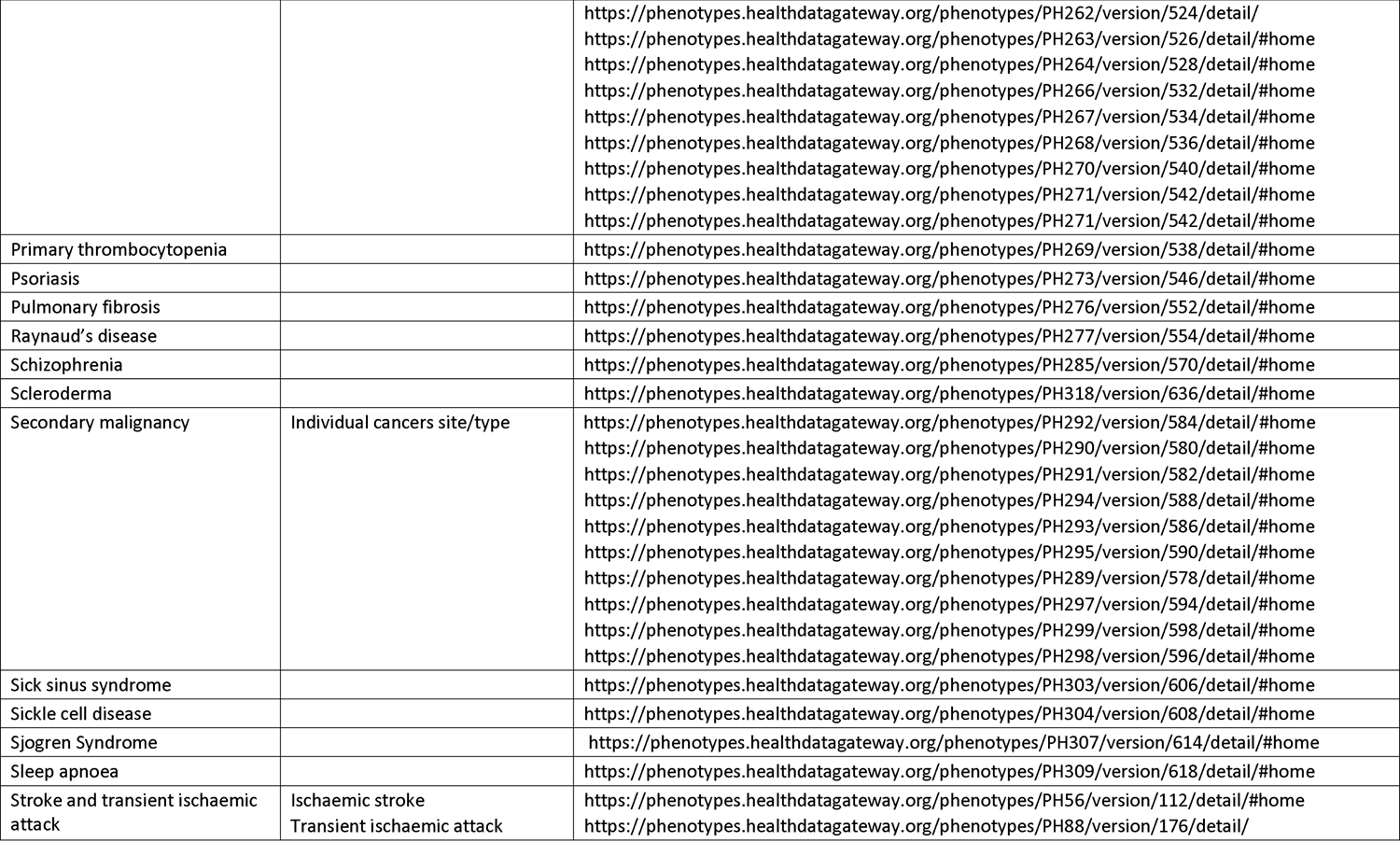
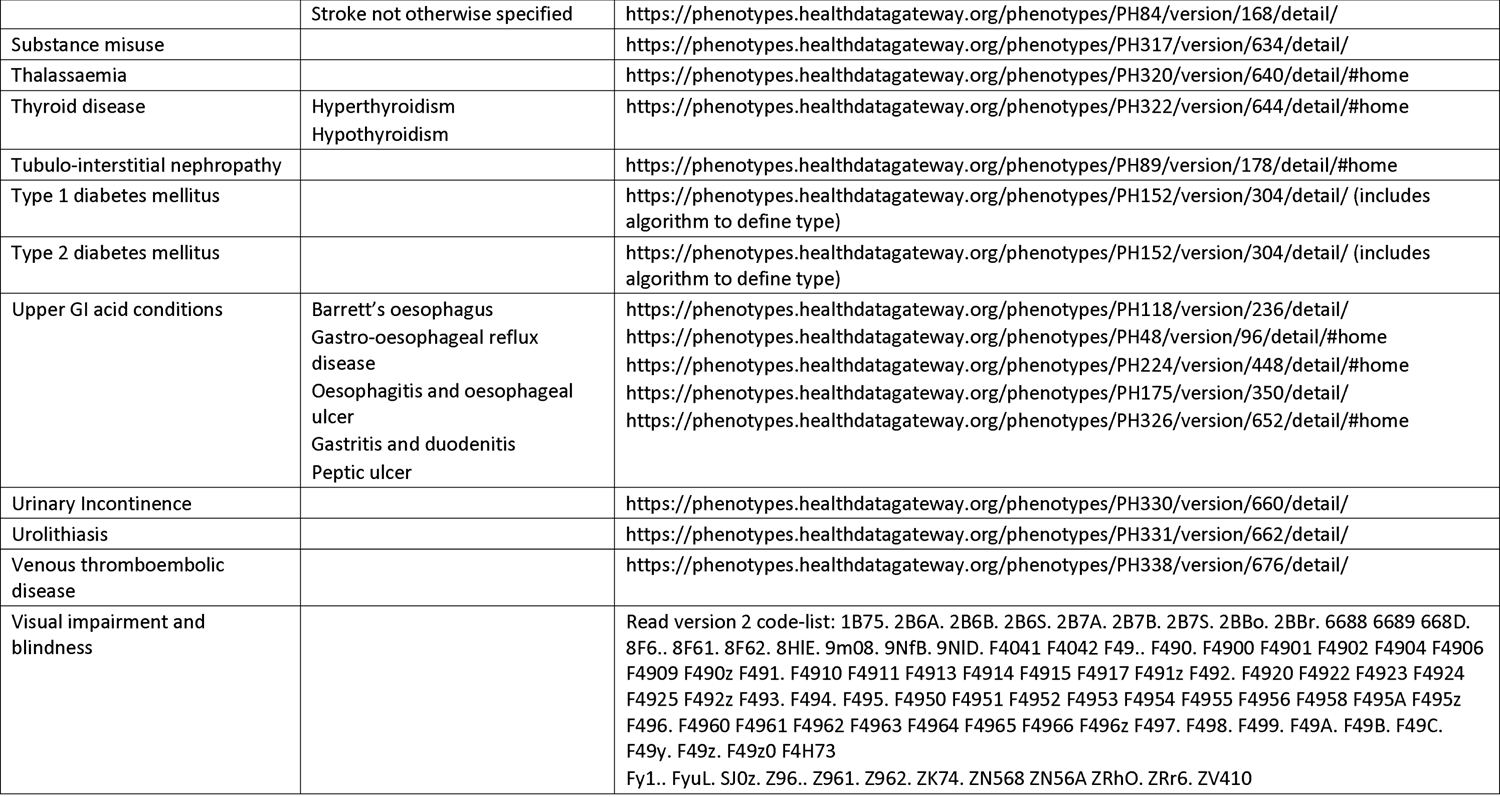
List of chronic conditions examined, and codes used to define those morbidities

| Condition used in this analysis | Condition subsets (to map to code lists) | Codes used or code list source |
| --- | --- | --- |
| Addison's Disease |  | Read v2 codes A176. C154. C1540 C1541 C1546 C154z<br>ICD10 codes E27.1 E27.2 |
| Alcohol misuse |  | https://phenotypes.healthdatagateway.org/phenotypes/PH94/version/188/detail/#home |
| Ankylosing spondylitis |  | https://phenotypes.healthdatagateway.org/phenotypes/PH99/version/198/detail/#home |
| Anxiety |  | https://phenotypes.healthdatagateway.org/phenotypes/PH104/version/208/detail/#home |
| Aplastic anaemia |  | https://phenotypes.healthdatagateway.org/phenotypes/PH105/version/210/detail/#home |
| Asthma |  | https://phenotypes.healthdatagateway.org/phenotypes/PH109/version/218/detail/#home |
| Atrial fibrillation |  | https://phenotypes.healthdatagateway.org/phenotypes/PH36/version/72/detail/#home |
| Autism |  | https://phenotypes.healthdatagateway.org/phenotypes/PH110/version/220/detail/#home |
| Benign prostatic hyperplasia |  | https://phenotypes.healthdatagateway.org/phenotypes/PH39/version/78/detail/#home |
| Bipolar affective disorder |  | https://phenotypes.healthdatagateway.org/phenotypes/PH38/version/76/detail/#home |
| Bronchiectasis |  | https://phenotypes.healthdatagateway.org/phenotypes/PH128/version/256/detail/#home |
| Cardiomyopathy | Dilated cardiomyopathy<br>Hypertrophic cardiomyopathy<br>Cardiomyopathy – other | https://phenotypes.healthdatagateway.org/phenotypes/PH145/version/290/detail/#home<br>https://phenotypes.healthdatagateway.org/phenotypes/PH185/version/370/detail/#home<br>https://phenotypes.healthdatagateway.org/phenotypes/PH129/version/258/detail/#home |
| Cerebral palsy |  | https://phenotypes.healthdatagateway.org/phenotypes/PH132/version/264/detail/#home |
| Chronic kidney disease | Chronic kidney disease (laboratory result defined)<br>End stage renal disease | https://phenotypes.healthdatagateway.org/phenotypes/PH42/version/1527/detail/#home<br>https://phenotypes.healthdatagateway.org/phenotypes/PH45/version/90/detail/#home |
| Chronic obstructive pulmonary disease |  | https://phenotypes.healthdatagateway.org/phenotypes/PH43/version/86/detail/#home |
| Coeliac disease |  | https://phenotypes.healthdatagateway.org/phenotypes/PH141/version/282/detail/#home |
| Coronary heart disease (CHD) | Myocardial infarction<br>Stable angina<br>Unstable angina | https://phenotypes.healthdatagateway.org/phenotypes/PH215/version/430/detail/#home<br>https://phenotypes.healthdatagateway.org/phenotypes/PH315/version/630/detail/#home<br>https://phenotypes.healthdatagateway.org/phenotypes/PH329/version/658/detail/#home |
|  | CHD not otherwise specified | https://phenotypes.healthdatagateway.org/phenotypes/PH41/version/82/detail/#home |
| Cystic fibrosis |  | https://phenotypes.healthdatagateway.org/phenotypes/PH40/version/80/detail/#home |
| Dementia |  | https://phenotypes.healthdatagateway.org/phenotypes/PH148/version/296/detail/#home |
| Depression |  | https://phenotypes.healthdatagateway.org/phenotypes/PH149/version/298/detail/#home |
| Diabetes mellitus other or not specified |  | https://phenotypes.healthdatagateway.org/phenotypes/PH152/version/304/detail/ |
| Diverticular disease |  | https://phenotypes.healthdatagateway.org/phenotypes/PH154/version/308/detail/#home |
| Eating disorders |  | https://phenotypes.healthdatagateway.org/phenotypes/PH159/version/318/detail/#home |
| Endometriosis |  | https://phenotypes.healthdatagateway.org/phenotypes/PH162/version/324/detail/ |
| Epilepsy |  | https://phenotypes.healthdatagateway.org/phenotypes/PH165/version/330/detail/#home |
| Erectile dysfunction |  | https://phenotypes.healthdatagateway.org/phenotypes/PH44/version/88/detail/#home |
| Giant cell arteritis |  | https://phenotypes.healthdatagateway.org/phenotypes/PH46/version/92/detail/#home |
| Gout |  | https://phenotypes.healthdatagateway.org/phenotypes/PH177/version/354/detail/#home |
| Haematological malignancy |  | https://phenotypes.healthdatagateway.org/phenotypes/PH60/version/120/detail/#home<br>https://phenotypes.healthdatagateway.org/phenotypes/PH63/version/126/detail/#home<br>https://phenotypes.healthdatagateway.org/phenotypes/PH68/version/136/detail/#home<br>https://phenotypes.healthdatagateway.org/phenotypes/PH186/version/372/detail/<br>https://phenotypes.healthdatagateway.org/phenotypes/PH199/version/398/detail/<br>https://phenotypes.healthdatagateway.org/phenotypes/PH239/version/478/detail/#home |
| Hearing loss |  | Read version 2 codes 1C13300 2BL3.00 2BL4.00 F580100 F591400 F591600 F591C00 F592100 F598.00 1C17.00 2DG..00 2DH0.00 7311A00 7317C00 7320 7319600 7319 8D23.00 8D26.00 8D2..12 8M41.00 Z8B5300 Z8B5500 Z911100 Z911.00 8D2..12 ZV45G00 ZV45N00 ZV53200 ZV53D00 2BL5.00 F591900 F591A00 |
| Heart block and bundle branch block | Atrioventricular block second degree<br>Bifascicular block<br>Trifascicular block<br>Atrioventricular block third degree<br>Left bundle branch block<br>Right bundle branch block | https://phenotypes.healthdatagateway.org/phenotypes/PH114/version/228/detail/<br>https://phenotypes.healthdatagateway.org/phenotypes/PH125/version/250/detail/<br>https://phenotypes.healthdatagateway.org/phenotypes/PH324/version/648/detail/#home<br>https://phenotypes.healthdatagateway.org/phenotypes/PH115/version/230/detail/<br>https://phenotypes.healthdatagateway.org/phenotypes/PH57/version/114/detail/#home<br>https://phenotypes.healthdatagateway.org/phenotypes/PH78/version/156/detail/#home |
| Heart failure |  | https://phenotypes.healthdatagateway.org/phenotypes/PH182/version/364/detail/#home |
| Heart valve disease | Rheumatic valve disorder<br>Non-rheumatic aortic valve disorder<br>Non-rheumatic mitral valve disorder<br>Multiple valve disorder | https://phenotypes.healthdatagateway.org/phenotypes/PH81/version/162/detail/<br>https://phenotypes.healthdatagateway.org/phenotypes/PH219/version/438/detail/#home<br>https://phenotypes.healthdatagateway.org/phenotypes/PH220/version/440/detail/#home<br>https://phenotypes.healthdatagateway.org/phenotypes/PH212/version/424/detail/#home |
| Human immunodeficiency virus |  | https://phenotypes.healthdatagateway.org/phenotypes/PH184/version/368/detail/#home |
| Hyperparathyroidism |  | https://phenotypes.healthdatagateway.org/phenotypes/PH76/version/152/detail/ |
| Hypertension |  | https://phenotypes.healthdatagateway.org/phenotypes/PH189/version/378/detail/#home |
| Inflammatory arthritis | Juvenile Arthritis<br>Psoriatic Arthritis<br>Reactive Arthritis<br>Rheumatoid Arthritis | https://phenotypes.healthdatagateway.org/phenotypes/PH196/version/392/detail/<br>https://phenotypes.healthdatagateway.org/phenotypes/PH80/version/160/detail/ |
| Inflammatory bowel disease | Crohn's Disease<br>Ulcerative colitis | https://phenotypes.healthdatagateway.org/phenotypes/PH144/version/288/detail/<br>https://phenotypes.healthdatagateway.org/phenotypes/PH326/version/652/detail/#home |
| Intellectual disability | Down Syndrome<br>Other intellectual disability | https://phenotypes.healthdatagateway.org/phenotypes/PH156/version/312/detail/<br>https://phenotypes.healthdatagateway.org/phenotypes/PH192/version/384/detail/ |
| Liver disease | Alcoholic liver disease<br>Autoimmune liver disease<br>Chronic viral hepatitis<br>Cirrhosis and liver fibrosis<br>Fatty liver<br>Liver failure<br>Oesophageal varices<br>Portal hypertension | https://phenotypes.healthdatagateway.org/phenotypes/PH202/version/404/detail/<br>https://phenotypes.healthdatagateway.org/phenotypes/PH111/version/222/detail/<br>https://phenotypes.healthdatagateway.org/phenotypes/PH137/version/274/detail/<br>https://phenotypes.healthdatagateway.org/phenotypes/PH140/version/280/detail/<br>https://phenotypes.healthdatagateway.org/phenotypes/PH168/version/336/detail/<br>https://phenotypes.healthdatagateway.org/phenotypes/PH203/version/406/detail/<br>https://phenotypes.healthdatagateway.org/phenotypes/PH334/version/668/detail/#home<br>https://phenotypes.healthdatagateway.org/phenotypes/PH243/version/486/detail/#home |
| Lupus Erythematosis |  | https://phenotypes.healthdatagateway.org/phenotypes/PH83/version/166/detail/#home |
| Motor Neurone Disease |  | https://phenotypes.healthdatagateway.org/phenotypes/PH62/version/124/detail/#home |
| Multiple sclerosis |  | https://phenotypes.healthdatagateway.org/phenotypes/PH63/version/126/detail/#home |
| Myasthenia gravis |  | https://phenotypes.healthdatagateway.org/phenotypes/PH213/version/426/detail/#home |
| Neuropathic bladder |  | https://phenotypes.healthdatagateway.org/phenotypes/PH218/version/436/detail/#home |
| Obsessive compulsive disorder |  | https://phenotypes.healthdatagateway.org/phenotypes/PH223/version/446/detail/#home |
| Obstructive and reflux uropathy |  | https://phenotypes.healthdatagateway.org/phenotypes/PH222/version/444/detail/ |
| Osteoarthritis |  | https://phenotypes.healthdatagateway.org/phenotypes/PH63/version/126/detail/#home |
| Osteoporosis |  | https://phenotypes.healthdatagateway.org/phenotypes/PH225/version/450/detail/#home |
| Paralysis |  | Read codes version 2: 2835 F241100 F141.00 F241.00 F241000 F230000 F240.11 F240100 F240000 F232.11 F232.00 2836 F240.00 F242.00 F230.00 F230z00 2837 F230100 |
| Parkinson's Disease |  | https://phenotypes.healthdatagateway.org/phenotypes/PH77/version/154/detail/ |
| Paroxysmal tachycardias | Supraventricular tachycardia<br>Ventricular tachycardia | https://phenotypes.healthdatagateway.org/phenotypes/PH84/version/168/detail/#home<br>https://phenotypes.healthdatagateway.org/phenotypes/PH90/version/180/detail/#home |
| Peripheral artery disease |  | https://phenotypes.healthdatagateway.org/phenotypes/PH235/version/470/detail/#home |
| Peripheral neuropathy | Peripheral neuropathy<br>Autonomic neuropathy | https://phenotypes.healthdatagateway.org/phenotypes/PH235/version/470/detail/#home<br>https://phenotypes.healthdatagateway.org/phenotypes/PH112/version/224/detail/<br>https://phenotypes.healthdatagateway.org/phenotypes/PH155/version/310/detail/#home<br>(selected codes) |
| Polymyalgia rheumatica |  | https://phenotypes.healthdatagateway.org/phenotypes/PH74/version/148/detail/#home |
| Primary malignancy | Individual cancers site/type | https://phenotypes.healthdatagateway.org/phenotypes/PH250/version/500/detail/#home<br>https://phenotypes.healthdatagateway.org/phenotypes/PH252/version/504/detail/#home<br>https://phenotypes.healthdatagateway.org/phenotypes/PH256/version/512/detail/#home<br>https://phenotypes.healthdatagateway.org/phenotypes/PH265/version/530/detail/#home<br>https://phenotypes.healthdatagateway.org/phenotypes/PH247/version/494/detail/#home<br>https://phenotypes.healthdatagateway.org/phenotypes/PH248/version/496/detail/#home<br>https://phenotypes.healthdatagateway.org/phenotypes/PH249/version/498/detail/#home<br>https://phenotypes.healthdatagateway.org/phenotypes/PH251/version/502/detail/#home<br>https://phenotypes.healthdatagateway.org/phenotypes/PH253/version/506/detail/#home<br>https://phenotypes.healthdatagateway.org/phenotypes/PH254/version/508/detail/#home<br>https://phenotypes.healthdatagateway.org/phenotypes/PH255/version/510/detail/#home<br>https://phenotypes.healthdatagateway.org/phenotypes/PH257/version/514/detail/#home<br>https://phenotypes.healthdatagateway.org/phenotypes/PH258/version/516/detail/#home<br>https://phenotypes.healthdatagateway.org/phenotypes/PH259/version/518/detail/#home<br>https://phenotypes.healthdatagateway.org/phenotypes/PH260/version/520/detail/#home<br>https://phenotypes.healthdatagateway.org/phenotypes/PH260/version/520/detail/#home |
|  |  | https://phenotypes.healthdatagateway.org/phenotypes/PH262/version/524/detail/<br>https://phenotypes.healthdatagateway.org/phenotypes/PH263/version/526/detail/#home<br>https://phenotypes.healthdatagateway.org/phenotypes/PH264/version/528/detail/#home<br>https://phenotypes.healthdatagateway.org/phenotypes/PH266/version/532/detail/#home<br>https://phenotypes.healthdatagateway.org/phenotypes/PH267/version/534/detail/#home<br>https://phenotypes.healthdatagateway.org/phenotypes/PH268/version/536/detail/#home<br>https://phenotypes.healthdatagateway.org/phenotypes/PH270/version/540/detail/#home<br>https://phenotypes.healthdatagateway.org/phenotypes/PH271/version/542/detail/#home<br>https://phenotypes.healthdatagateway.org/phenotypes/PH271/version/542/detail/#home |
| Primary thrombocytopenia |  | https://phenotypes.healthdatagateway.org/phenotypes/PH269/version/538/detail/#home |
| Psoriasis |  | https://phenotypes.healthdatagateway.org/phenotypes/PH273/version/546/detail/#home |
| Pulmonary fibrosis |  | https://phenotypes.healthdatagateway.org/phenotypes/PH276/version/552/detail/#home |
| Raynaud's disease |  | https://phenotypes.healthdatagateway.org/phenotypes/PH277/version/554/detail/#home |
| Schizophrenia |  | https://phenotypes.healthdatagateway.org/phenotypes/PH285/version/570/detail/#home |
| Scleroderma |  | https://phenotypes.healthdatagateway.org/phenotypes/PH318/version/636/detail/#home |
| Secondary malignancy | Individual cancers site/type | https://phenotypes.healthdatagateway.org/phenotypes/PH292/version/584/detail/#home<br>https://phenotypes.healthdatagateway.org/phenotypes/PH290/version/580/detail/#home<br>https://phenotypes.healthdatagateway.org/phenotypes/PH291/version/582/detail/#home<br>https://phenotypes.healthdatagateway.org/phenotypes/PH294/version/588/detail/#home<br>https://phenotypes.healthdatagateway.org/phenotypes/PH293/version/586/detail/#home<br>https://phenotypes.healthdatagateway.org/phenotypes/PH295/version/590/detail/#home<br>https://phenotypes.healthdatagateway.org/phenotypes/PH289/version/578/detail/#home<br>https://phenotypes.healthdatagateway.org/phenotypes/PH297/version/594/detail/#home<br>https://phenotypes.healthdatagateway.org/phenotypes/PH299/version/598/detail/#home<br>https://phenotypes.healthdatagateway.org/phenotypes/PH298/version/596/detail/#home |
| Sick sinus syndrome |  | https://phenotypes.healthdatagateway.org/phenotypes/PH303/version/606/detail/#home |
| Sickle cell disease |  | https://phenotypes.healthdatagateway.org/phenotypes/PH304/version/608/detail/#home |
| Sjogren Syndrome |  | https://phenotypes.healthdatagateway.org/phenotypes/PH307/version/614/detail/#home |
| Sleep apnoea |  | https://phenotypes.healthdatagateway.org/phenotypes/PH309/version/618/detail/#home |
| Stroke and transient ischaemic attack | Ischaemic stroke<br>Transient ischaemic attack | https://phenotypes.healthdatagateway.org/phenotypes/PH56/version/112/detail/#home<br>https://phenotypes.healthdatagateway.org/phenotypes/PH88/version/176/detail/ |
|  | Stroke not otherwise specified | https://phenotypes.healthdatagateway.org/phenotypes/PH84/version/168/detail/ |
| Substance misuse |  | https://phenotypes.healthdatagateway.org/phenotypes/PH317/version/634/detail/ |
| Thalassaemia |  | https://phenotypes.healthdatagateway.org/phenotypes/PH320/version/640/detail/#home |
| Thyroid disease | Hyperthyroidism<br>Hypothyroidism | https://phenotypes.healthdatagateway.org/phenotypes/PH322/version/644/detail/#home |
| Tubulo-interstitial nephropathy |  | https://phenotypes.healthdatagateway.org/phenotypes/PH89/version/178/detail/#home |
| Type 1 diabetes mellitus |  | https://phenotypes.healthdatagateway.org/phenotypes/PH152/version/304/detail/ (includes algorithm to define type) |
| Type 2 diabetes mellitus |  | https://phenotypes.healthdatagateway.org/phenotypes/PH152/version/304/detail/ (includes algorithm to define type) |
| Upper GI acid conditions | Barrett's oesophagus<br>Gastro-oesophageal reflux disease<br>Oesophagitis and oesophageal ulcer<br>Gastritis and duodenitis<br>Peptic ulcer | https://phenotypes.healthdatagateway.org/phenotypes/PH118/version/236/detail/<br>https://phenotypes.healthdatagateway.org/phenotypes/PH48/version/96/detail/#home<br>https://phenotypes.healthdatagateway.org/phenotypes/PH224/version/448/detail/#home<br>https://phenotypes.healthdatagateway.org/phenotypes/PH175/version/350/detail/<br>https://phenotypes.healthdatagateway.org/phenotypes/PH326/version/652/detail/#home |
| Urinary Incontinence |  | https://phenotypes.healthdatagateway.org/phenotypes/PH330/version/660/detail/ |
| Urolithiasis |  | https://phenotypes.healthdatagateway.org/phenotypes/PH331/version/662/detail/ |
| Venous thromboembolic disease |  | https://phenotypes.healthdatagateway.org/phenotypes/PH338/version/676/detail/ |
| Visual impairment and blindness |  | Read version 2 code-list: 1B75. 2B6A. 2B6B. 2B6S. 2B7A. 2B7B. 2B7S. 2BBo. 2BBr. 6688 6689 668D. 8F6.. 8F61. 8F62. 8HIE. 9m08. 9NfB. 9NID. F4041 F4042 F49.. F490. F4900 F4901 F4902 F4904 F4906 F4909 F490z F491. F4910 F4911 F4913 F4914 F4915 F4917 F491z F492. F4920 F4922 F4923 F4924 F4925 F492z F493. F494. F495. F4950 F4951 F4952 F4953 F4954 F4955 F4956 F4958 F495A F495z F496. F4960 F4961 F4962 F4963 F4964 F4965 F4966 F496z F497. F498. F499. F49A. F49B. F49C. F49y. F49z. F49z0 F4H73<br>Fy1.. FyuL. SJ0z. Z96.. Z961. Z962. ZK74. ZN568 ZN56A ZRho. ZRr6. ZV410 |

**Supplementary Table 2.**
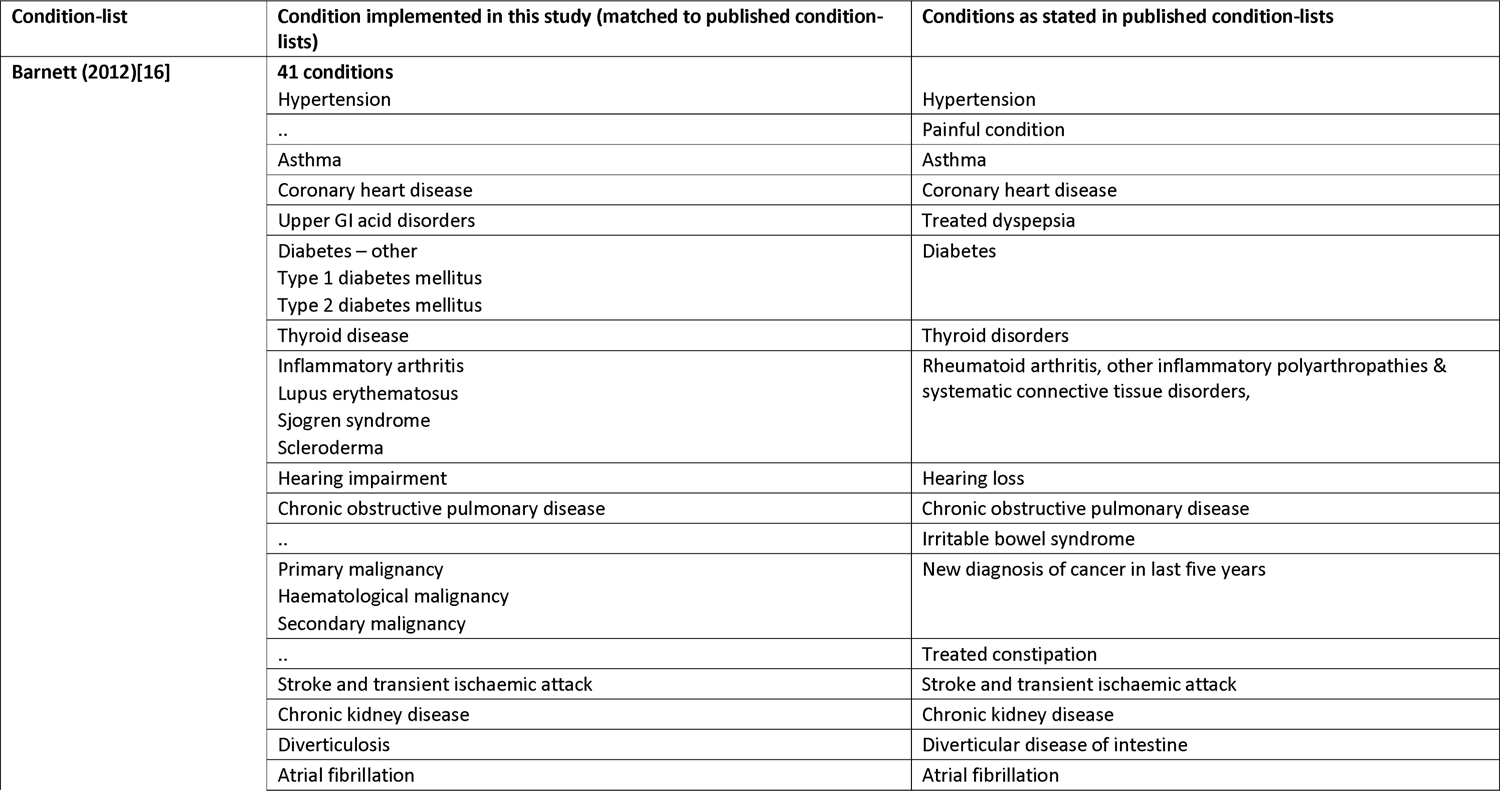
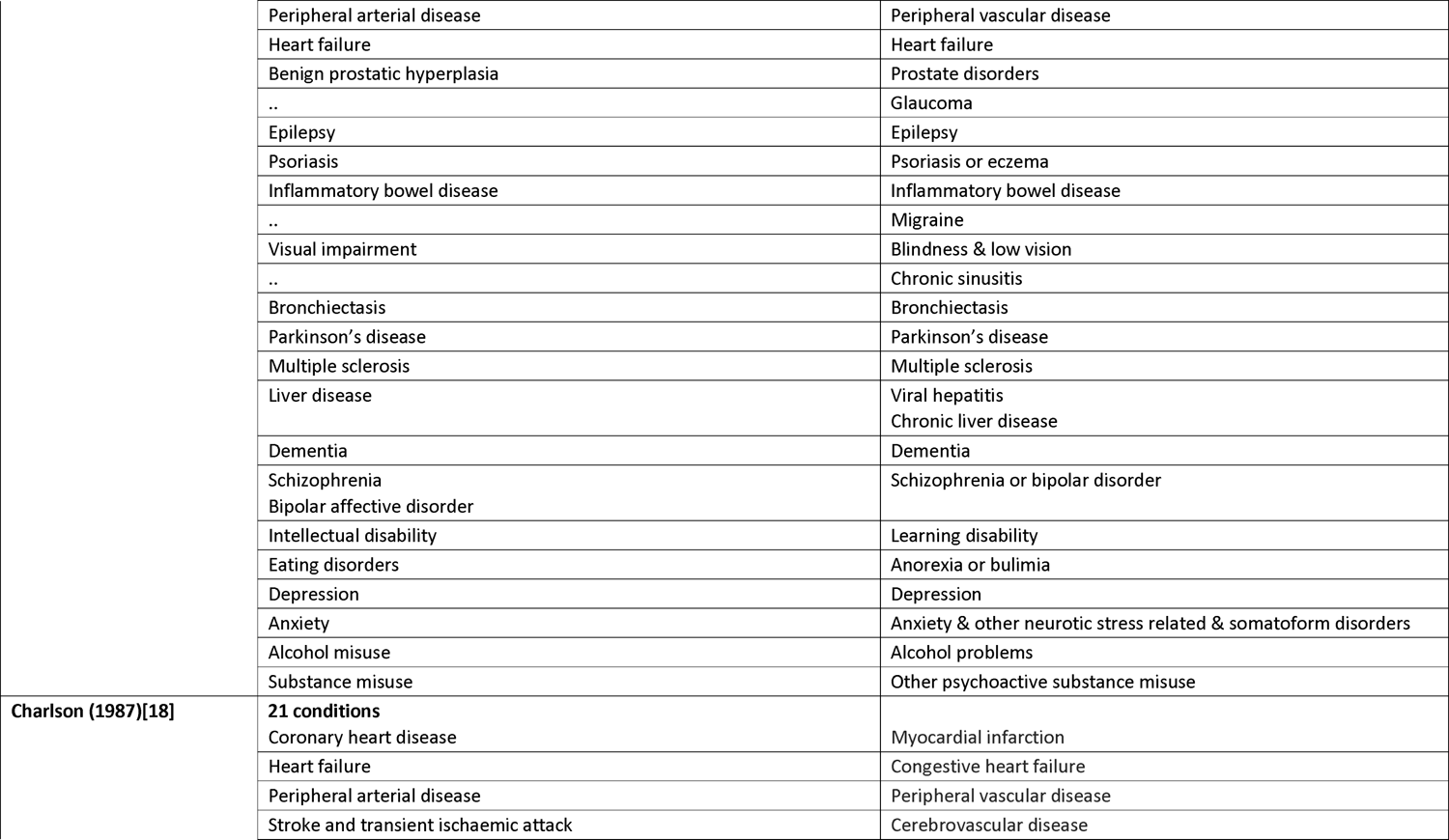
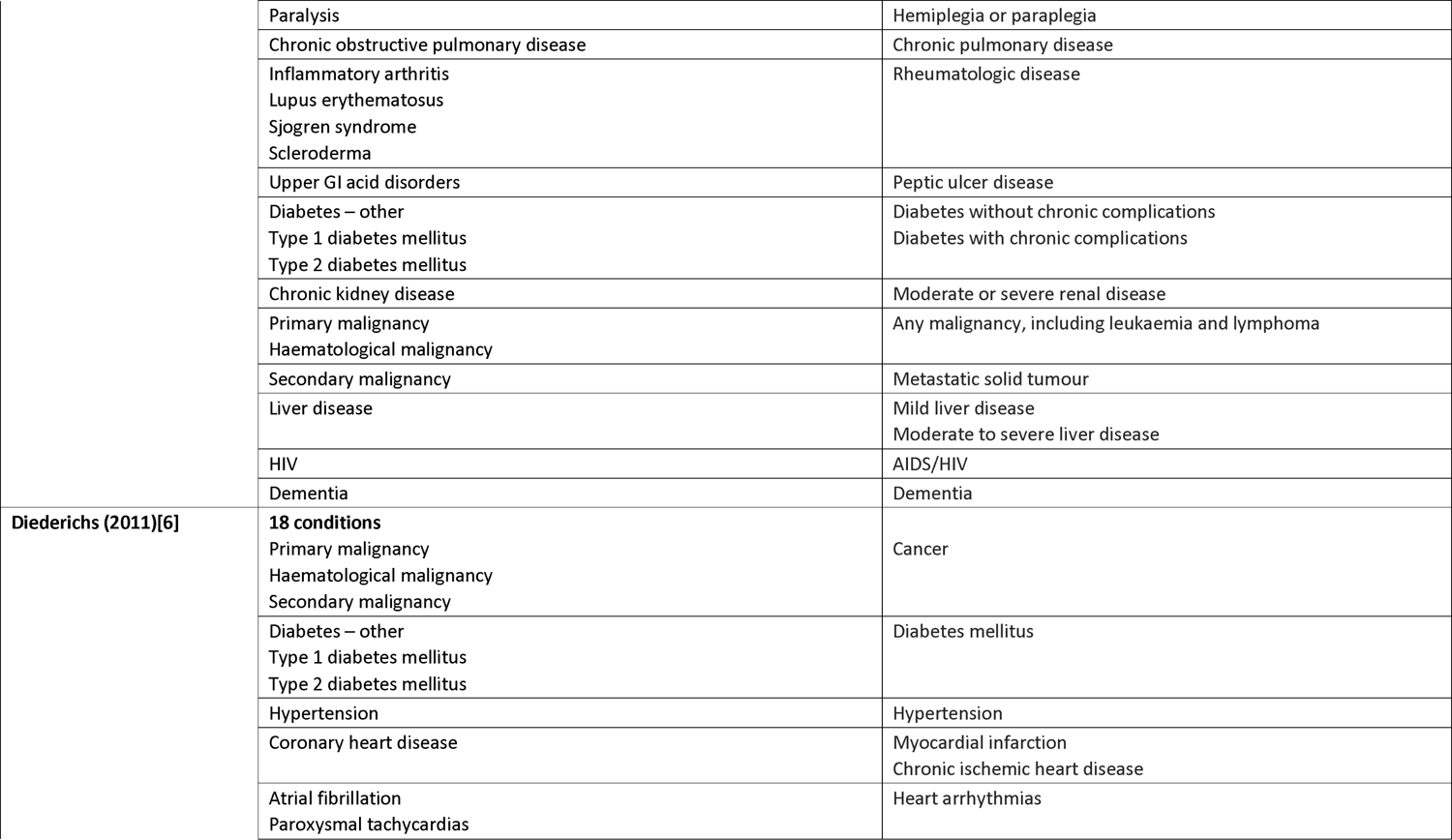
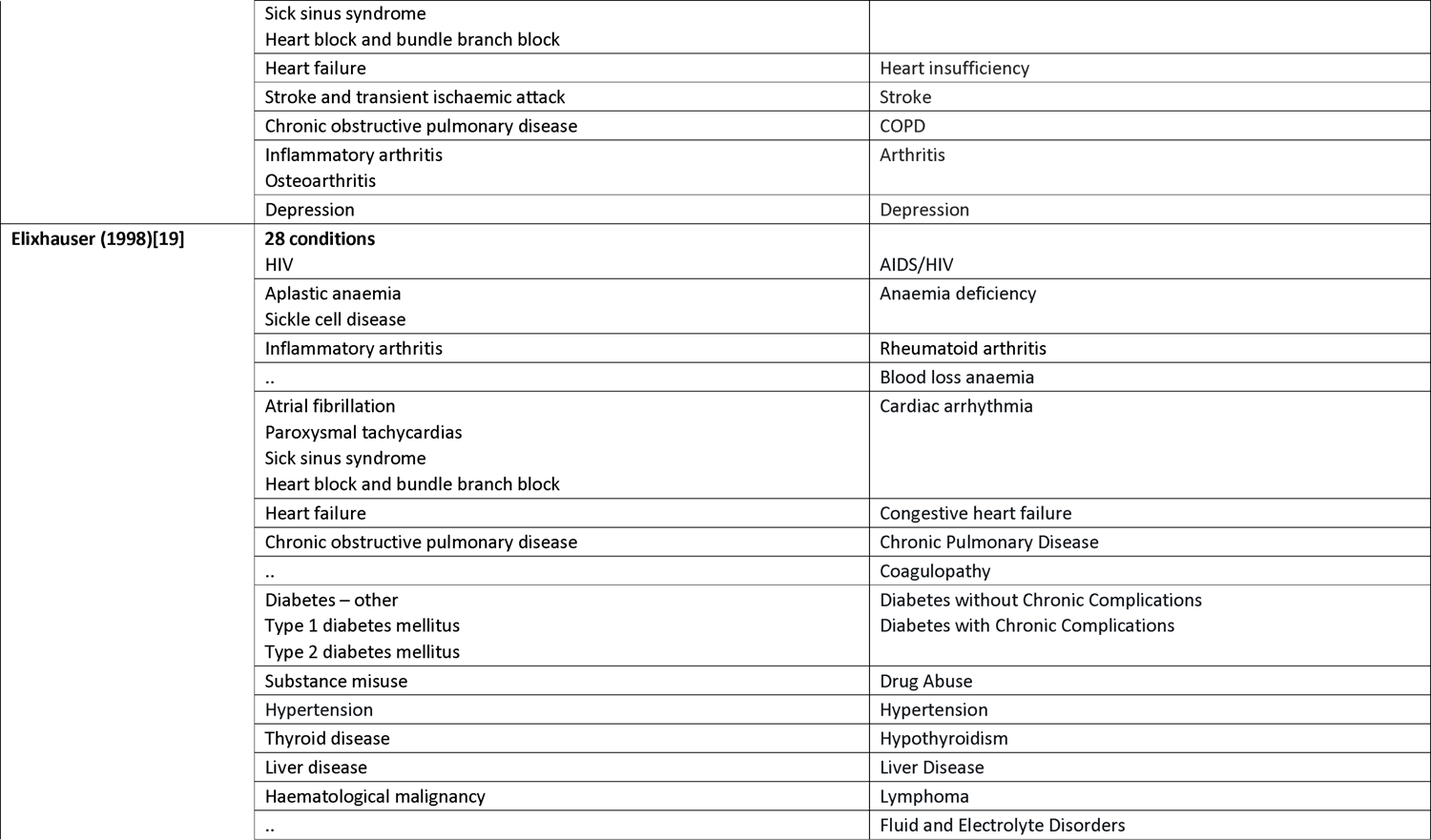
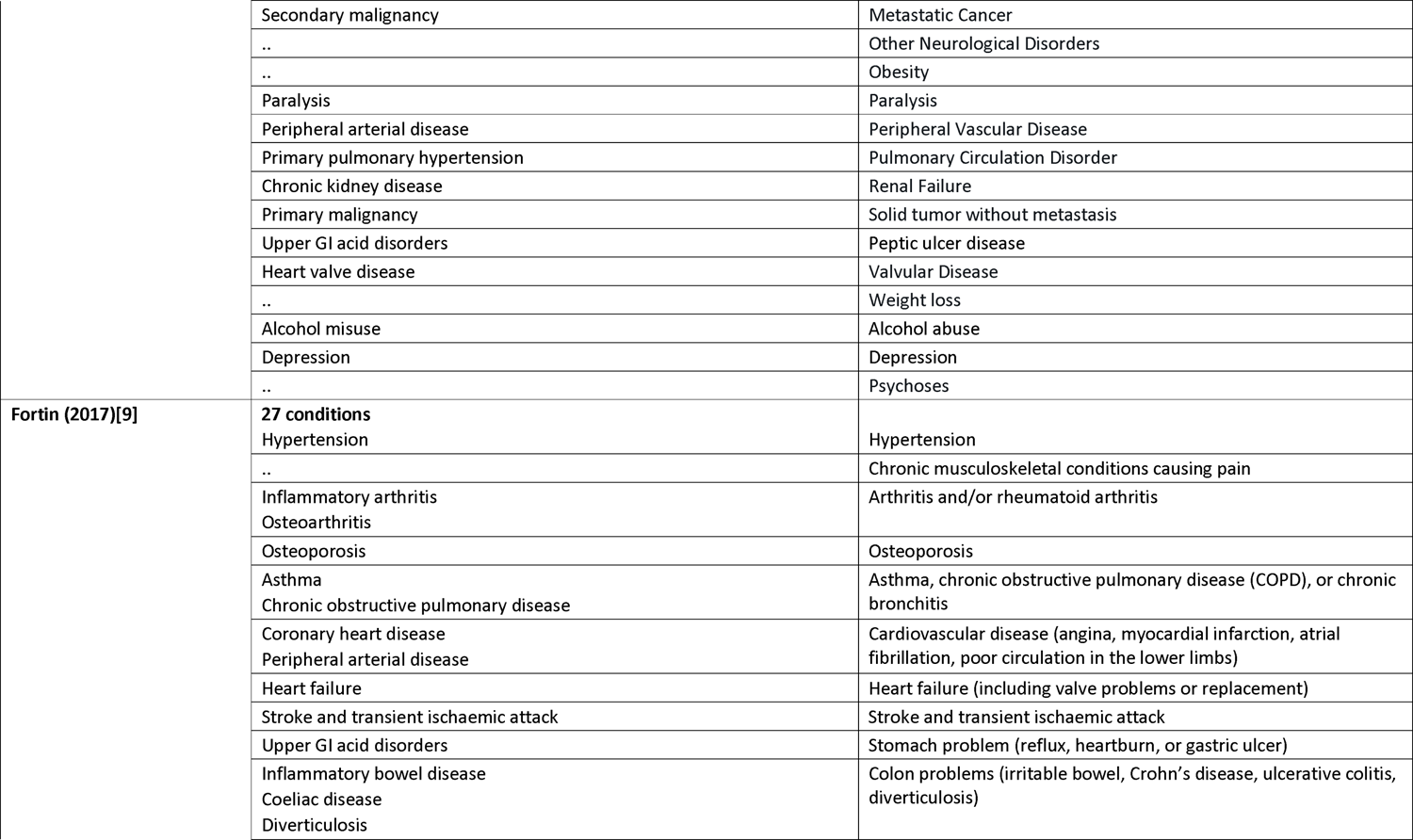
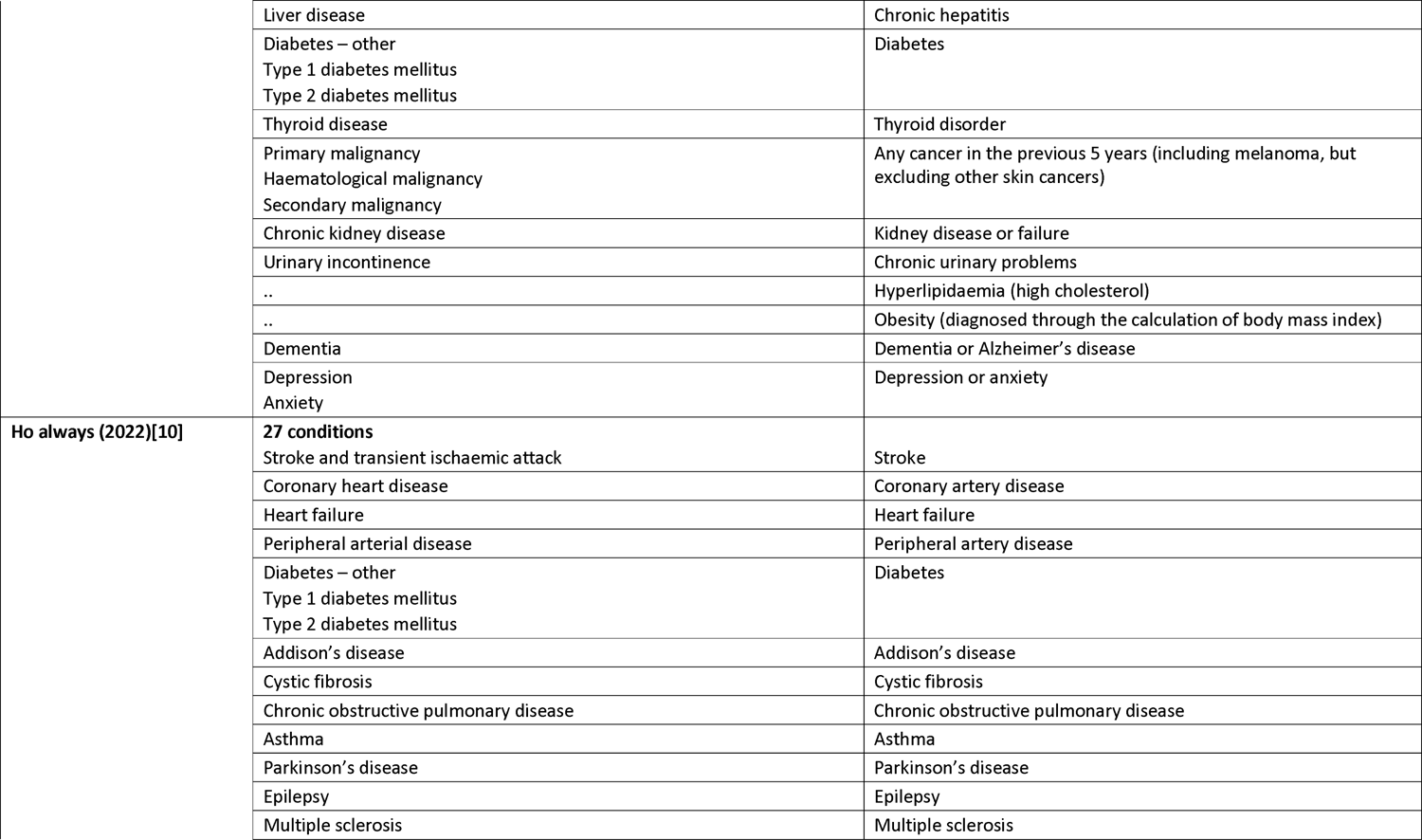
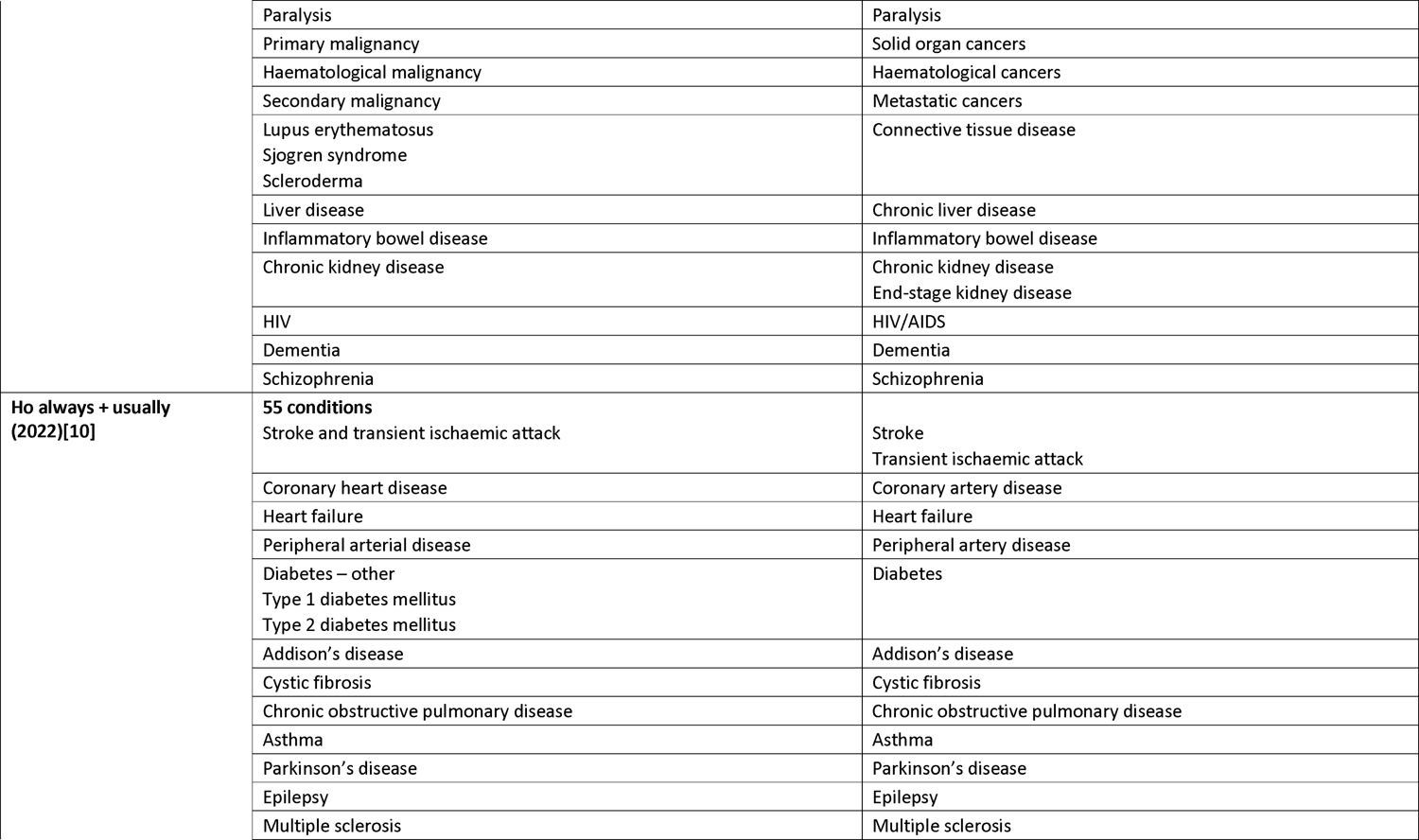
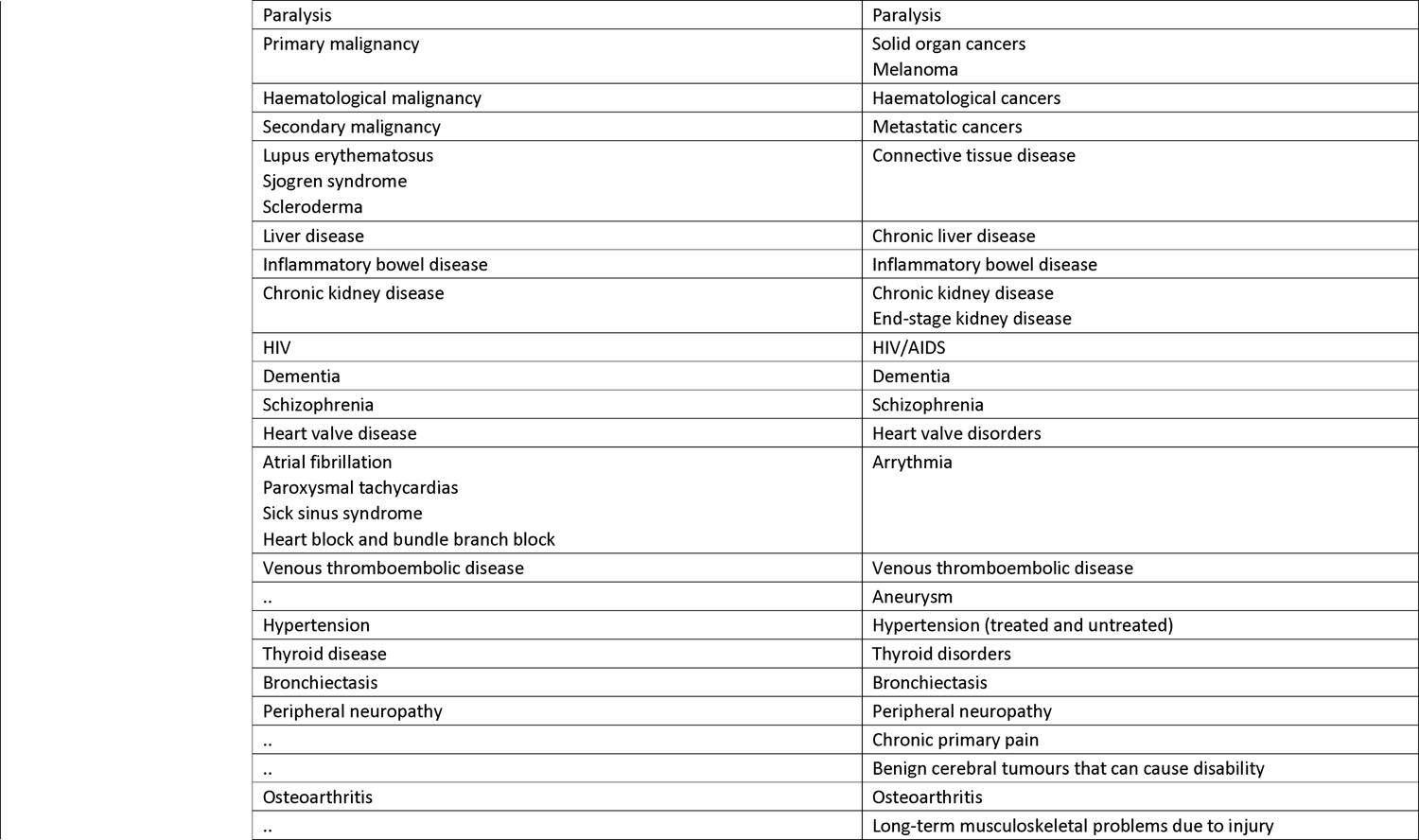
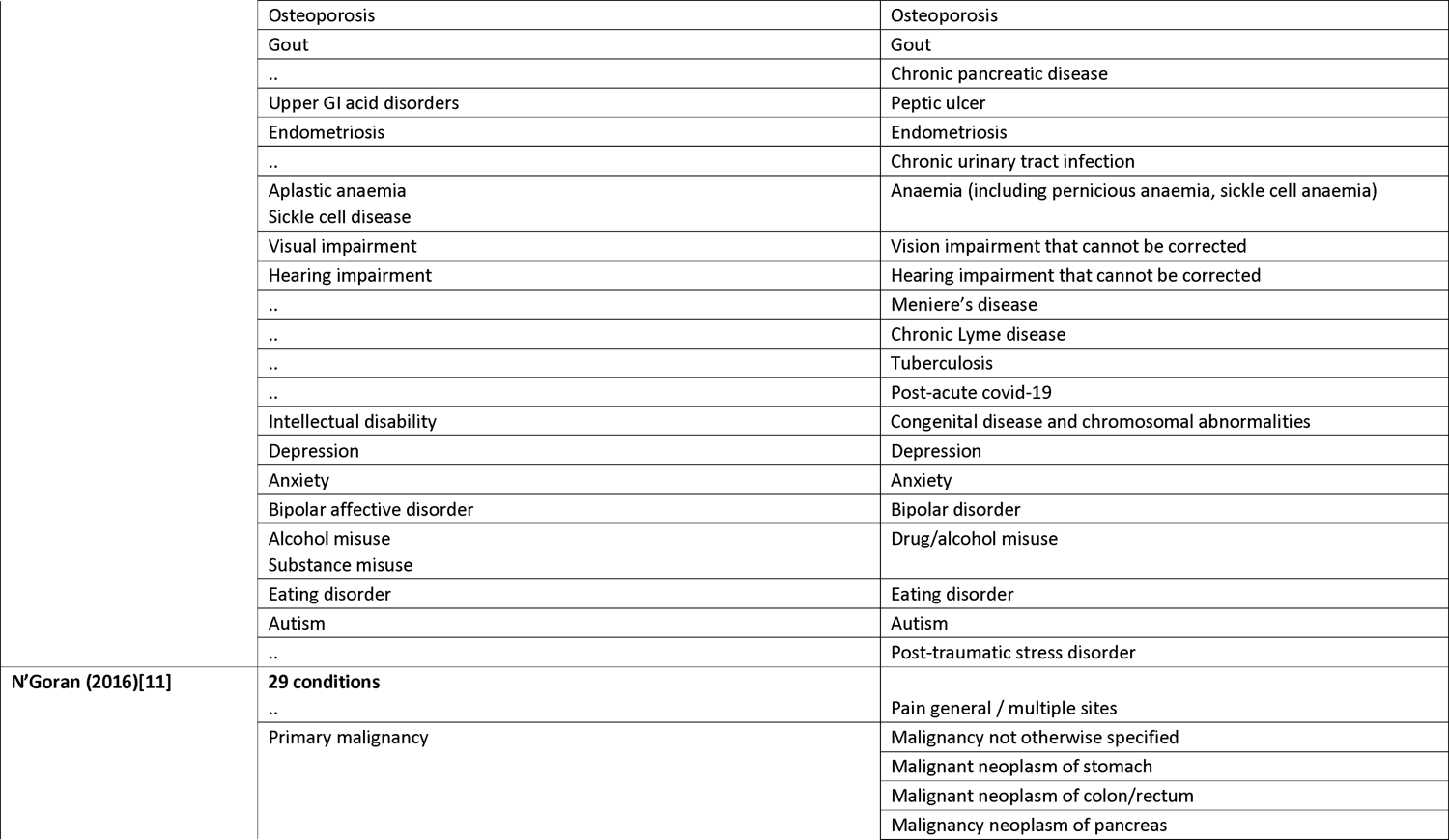
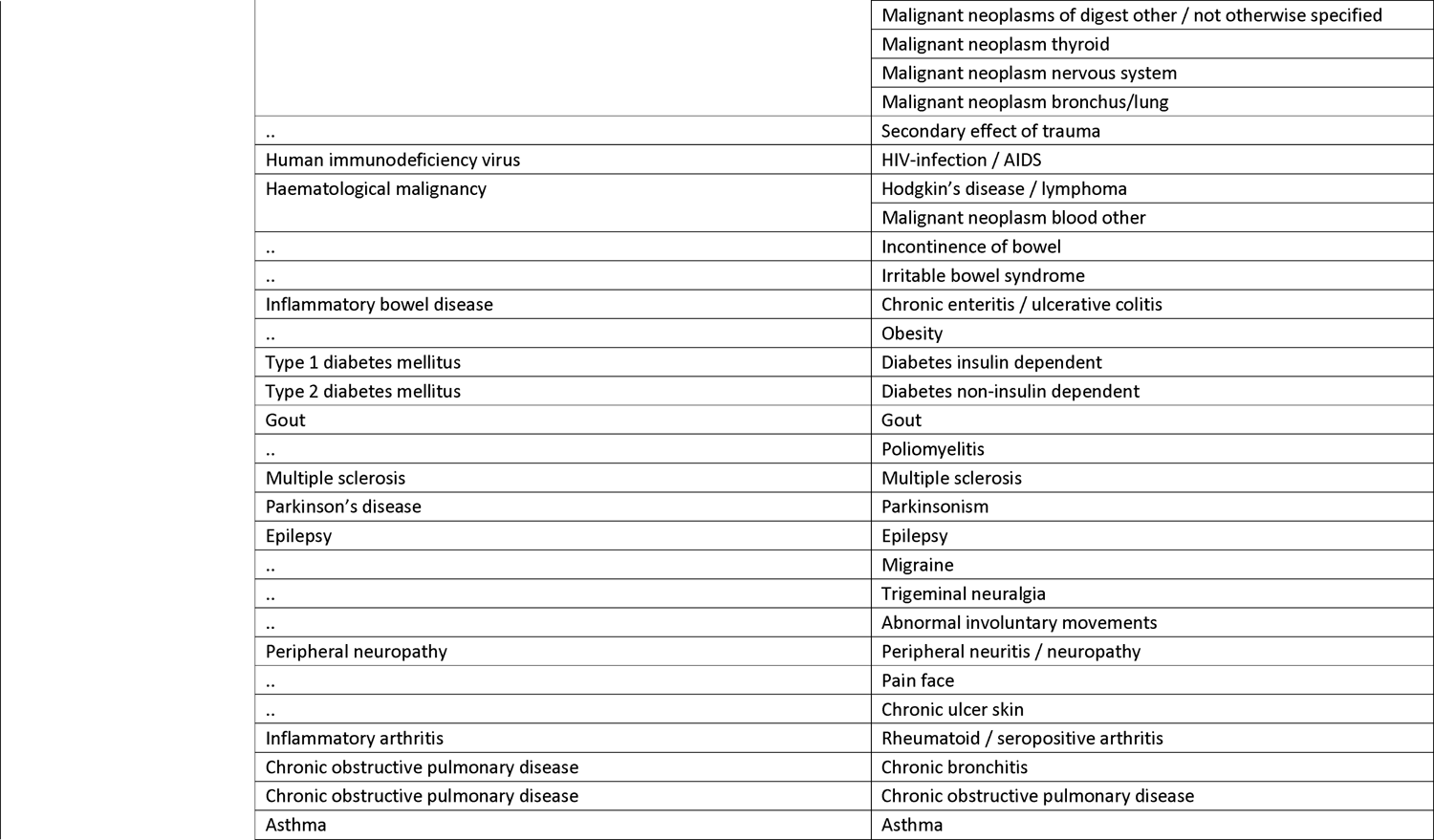
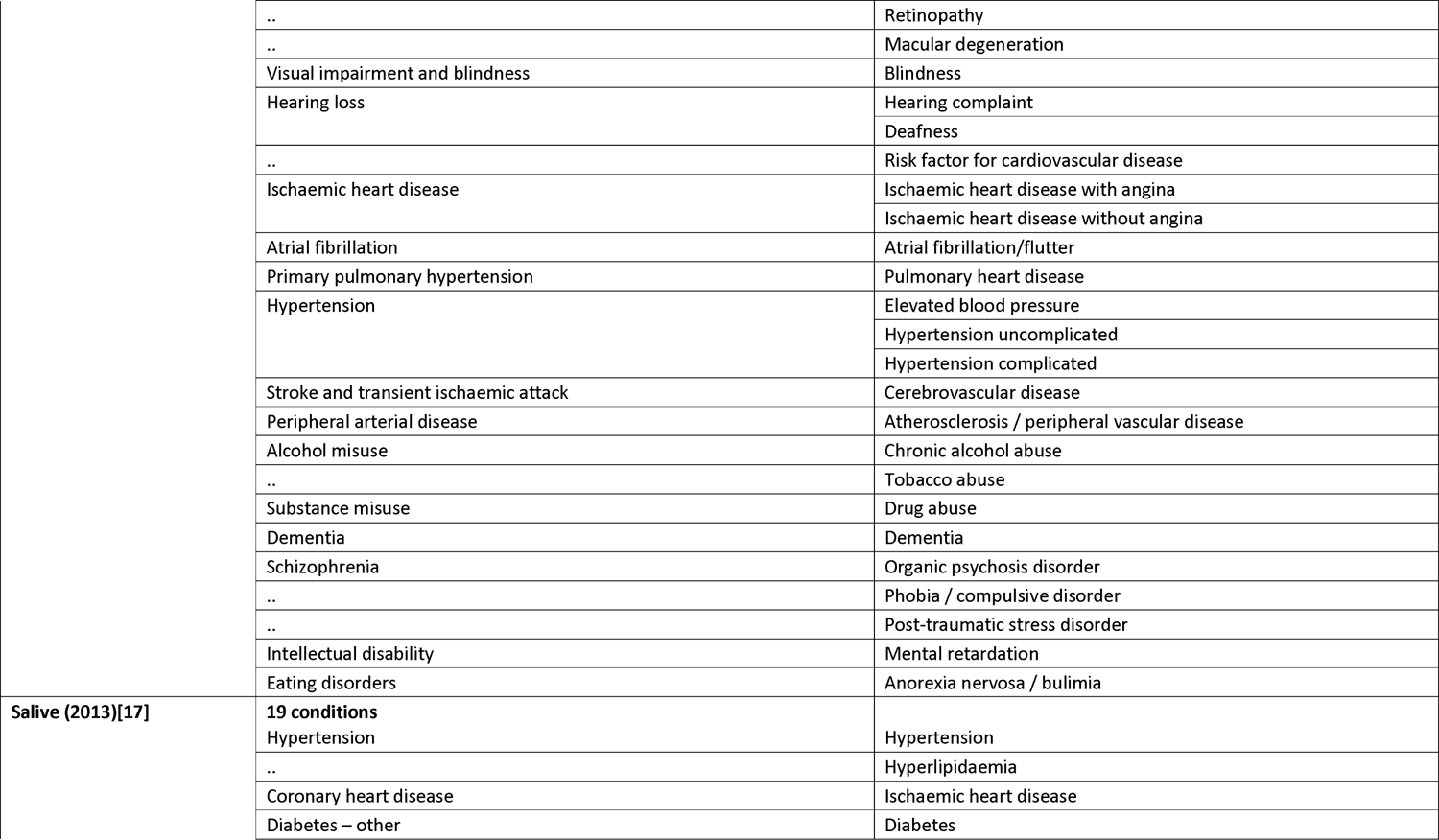
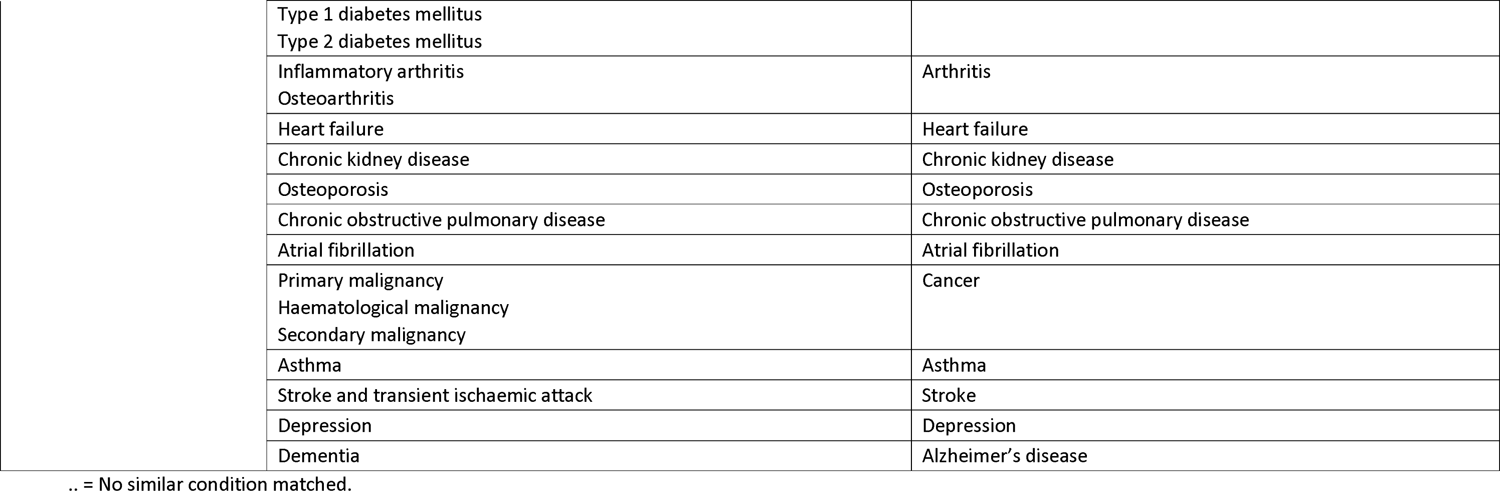
Conditions in each condition-list as implemented in this study, and as stated in published condition-lists.

**Supplementary Table 3.** The RECORD statement. Checklist of items, extended from the STROBE statement, that should be reported in observational studies using routinely collected health data.

|  | Item No. | STROBE items | Location in manuscript where items are reported | RECORD items | Location in manuscript where items are reported |
| --- | --- | --- | --- | --- | --- |
| **Title and abstract** |  |  |  |  |  |
|  | 1 | (a) Indicate the study's design with a commonly used term in the title or the abstract (b) Provide in the abstract an informative and balanced summary of what was done and what was found | **Title and abstract.**<br>**The study was a cross-sectional cohort study, and this is included in the abstract and methods sections.** | RECORD 1.1: The type of data used should be specified in the title or abstract. When possible, the name of the databases used should be included.<br>RECORD 1.2: If applicable, the geographic region and timeframe within which the study took place should be reported in the title or abstract.<br>RECORD 1.3: If linkage between databases was conducted for the study, this should be clearly stated in the title or abstract. | **1.1. Methods section of the abstract. The database name is used in the methods section of the main article.**<br>**1.2. Methods section of the abstract and main article.**<br>**1.3. Not applicable.** |
| **Introduction** |  |  |  |  |  |
| Background rationale | 2 | Explain the scientific background and rationale for the investigation being reported | **"Background" section of the abstract, and "Introduction" section of the main article.** |  |  |
| Objectives | 3 | State specific objectives, including any prespecified hypotheses | **Final paragraph of the "Background" section of the abstract, and "Introduction" section of the main article.** |  |  |
| **Methods** |  |  |  |  |  |
| Study Design | 4 | Present key elements of study design early in the paper | **“Study design” section of “Methods” in the main article.** |  |  |
| Setting | 5 | Describe the setting, locations, and relevant dates, including periods of recruitment, exposure, follow-up, and data collection | **“Methods” section of the abstract, and “Statistical analysis” section of “Methods” in the main article.** |  |  |
| Participants | 6 | *(a) Cohort study* - Give the eligibility criteria, and the sources and methods of selection of participants. Describe methods of follow-up<br>*Case-control study* - Give the eligibility criteria, and the sources and methods of case ascertainment and control selection. Give the rationale for the choice of cases and controls<br>*Cross-sectional study* - Give the eligibility criteria, and the sources and methods of selection of participants<br>*(b) Cohort study* - For matched studies, give matching criteria and number of exposed and unexposed<br>*Case-control study* - For matched studies, give matching criteria and the number of controls per case | **“Methods” section of the abstract, and “Results” section of the main article.** | RECORD 6.1: The methods of study population selection (such as codes or algorithms used to identify subjects) should be listed in detail. If this is not possible, an explanation should be provided.<br>RECORD 6.2: Any validation studies of the codes or algorithms used to select the population should be referenced. If validation was conducted for this study and not published elsewhere, detailed methods and results should be provided.<br>RECORD 6.3: If the study involved linkage of databases, consider use of a flow diagram or other graphical display to demonstrate the data linkage process, including the number of individuals with linked data at each stage. | **6.1 “Definition of variables” in “Methods” section of the main article explained the methodology we used to develop clinical code lists.**<br>**6.2 Code lists are available from the HDR UK Phenotype Library.**<br>**6.3 No data linkage was required.** |
| Variables | 7 | Clearly define all outcomes, exposures, predictors, potential confounders, and effect modifiers. Give diagnostic criteria, if applicable. | **“Statistical analysis” section of “Methods” in the main article.** | RECORD 7.1: A complete list of codes and algorithms used to classify exposures, outcomes, confounders, and effect modifiers should be provided. If these cannot be reported, an explanation should be provided. | **All code lists are available on the HDR UK Phenotype Library and listed in the supplementary material.** |
| Data sources/<br>measurement | 8 | For each variable of interest, give sources of data and details of methods of assessment (measurement). Describe comparability of assessment methods if there is more than one group | **“Data sources” and “Statistical analysis” sections of “Methods” in the main article.** |  |  |
| Bias | 9 | Describe any efforts to address potential sources of bias | **“Data sources” section of “Methods” in the main article reports how GP electronic health records, hospital admission data, and laboratory results were all used to define morbidities to combat bias where clinicians were not recording diagnoses.** |  |  |
| Study size | 10 | Explain how the study size was arrived at | **“Study participants” section of “Methods” in the main article. The entire population alive and registered with the 149 general practices at the study cross-section date were included. There was no need to take a sample.** |  |  |
| Quantitative variables | 11 | Explain how quantitative variables were handled in the analyses. If applicable, describe which groupings were chosen, and why | **Not applicable.** |  |  |
| Statistical methods | 12 | (a) Describe all statistical methods, including those used to control for confounding<br>(b) Describe any methods used to examine subgroups and interactions<br>(c) Explain how missing data were addressed<br>(d) *Cohort study* - If applicable, explain how loss to follow-up was addressed<br>*Case-control study* - If applicable, explain how matching of cases and controls was addressed<br>*Cross-sectional study* - If applicable, describe analytical methods taking account of sampling strategy<br>(e) Describe any sensitivity analyses | **“Statistical analysis” section of “Methods” in the main article.** |  |  |
| Data access and cleaning methods |  | .. |  | RECORD 12.1: Authors should describe the extent to which the investigators had access to the database population used to create the study population.<br>RECORD 12.2: Authors should provide information on the data cleaning methods used in the study.12.1 | **12.1 The last line of acknowledgements according to Lancet policy.**<br>**12.2 Minimal data cleaning was required due to the nature of the study.** |
| Linkage |  | .. |  | RECORD 12.3: State whether the study included person-level, institutional-level, or other data linkage across two or more databases. The methods of linkage and methods of linkage quality evaluation should be provided. | **No linkage was required.** |
| **Results** |  |  |  |  |  |
| Participants | 13 | (a) Report the numbers of individuals at each stage of the | **Not required as the whole population of** | RECORD 13.1: Describe in detail the selection of the persons included in the | **Not applicable as the whole population was** |
|  |  | study (*e.g.*, numbers potentially eligible, examined for eligibility, confirmed eligible, included in the study, completing follow-up, and analysed)<br>(b) Give reasons for non-participation at each stage.<br>(c) Consider use of a flow diagram | **the 149 general practices was included.** | study (*i.e.*, study population selection) including filtering based on data quality, data availability and linkage. The selection of included persons can be described in the text and/or by means of the study flow diagram. | **included. However, this is discussed in “Study participants” section of “Methods” in the main article.** |
| Descriptive data | 14 | (a) Give characteristics of study participants (*e.g.*, demographic, clinical, social) and information on exposures and potential confounders<br>(b) Indicate the number of participants with missing data for each variable of interest<br>(c) *Cohort study* - summarise follow-up time (*e.g.*, average and total amount) | **Table 1 main article.**<br>**No data were missing.** |  |  |
| Outcome data | 15 | *Cohort study* - Report numbers of outcome events or summary measures over time<br>*Case-control study* - Report numbers in each exposure category, or summary measures of exposure<br>*Cross-sectional study* - Report numbers of outcome events or summary measures | **Figures 1, 2, and 3 in the main article.** |  |  |
| Main results | 16 | (a) Give unadjusted estimates and, if applicable, confounder-adjusted estimates and their precision (*e.g.*, 95% confidence interval). Make clear which confounders were adjusted for and why they were included (b) Report category boundaries when continuous variables were categorized<br>(c) If relevant, consider translating estimates of relative risk into absolute risk for a meaningful time period | **“Results” section of the main article.** |  |  |
| Other analyses | 17 | Report other analyses done—e.g., analyses of subgroups and interactions, and sensitivity analyses | None undertaken. |  |  |
| **Discussion** |  |  |  |  |  |
| Key results | 18 | Summarise key results with reference to study objectives | **“Key findings” section and first paragraph of “Discussion” section of the main article.** |  |  |
| Limitations | 19 | Discuss limitations of the study, taking into account sources of potential bias or imprecision. Discuss both direction and magnitude of any potential bias | **“Discussion” section of the main article.** | RECORD 19.1: Discuss the implications of using data that were not created or collected to answer the specific research question(s). Include discussion of misclassification bias, unmeasured confounding, missing data, and changing eligibility over time, as they pertain to the study being reported. | **Section section of the second paragraph in the “Discussion” section of the main article.** |
| Interpretation | 20 | Give a cautious overall interpretation of results considering objectives, limitations, multiplicity of analyses, results from similar studies, and other relevant evidence | **“Discussion” section of the main article.** |  |  |
| Generalisability | 21 | Discuss the generalisability (external validity) of the study results | **“Discussion” section of the main article.** |  |  |
| **Other Information** |  |  |  |  |  |
| Funding | 22 | Give the source of funding and the role of the funders for the present study and, if applicable, for the original study on which the present article is based | **“Funding” section in the abstract.** |  |  |
| Accessibility of protocol, raw data, and programming code |  | .. |  | RECORD 22.1: Authors should provide information on how to access any supplemental information such as the study protocol, raw data, or programming code. | **Available in supplementary material.** |

\*Reference: Benchimol EI, Smeeth L, Guttman A, Harron K, Moher D, Petersen I, Sørensen HT, von Elm E, Langan SM, the RECORD Working Committee. The REporting of studies Conducted using Observational Routinely-collected health Data (RECORD) Statement. *PLoS Medicine* 2015; in press.
\*Checklist is protected under Creative Commons Attribution (CC BY) license

## References

1. Pefoyo AJK, Bronskill SE, Gruneir A, Calzavara A, Thavorn K, Petrosyan Y, et al. The increasing burden and complexity of multimorbidity. BMC Public Health. 2015;15(1):415-.

2. Violan C, Foguet-Boreu Q, Flores-Mateo G, Salisbury C, Blom J, Freitag M, et al. Prevalence, determinants and patterns of multimorbidity in primary care: a systematic review of observational studies. PloS one. 2014;9(7):e102149-e.

3. Multimorbidity: a priority for global health research. The Academy of Medical Sciences. Available at https://acmedsci.ac.uk/policy/policy-projects/multimorbidity [accessed 18 April 2022]

4. Ho IS-S, Azcoaga-Lorenzo A, Akbari A, Black C, Davies J, Hodgins P, et al. Examining variation in the measurement of multimorbidity in research: a systematic review of 566 studies. Lancet Public Health. 2021;6(8):e587–e97.

5. Marjorie C Johnston, Michael Crilly, Corri Black, Gordon J Prescott, Stewart W Mercer, Defining and measuring multimorbidity: a systematic review of systematic reviews, Eur J Public Health, Volume 29, Issue 1, February 2019, Pages 182–189, https://doi.org/10.1093/eurpub/cky098

6. Diederichs C, Berger K, Bartels DB. The measurement of multiple chronic diseases--a systematic review on existing multimorbidity indices. J Gerontol A Biol Sci Med Sci. 2011;66(3):301–11.

7. Ho IS-S, Azcoaga-Lorenzo A, Akbari A, Davies J, Hodgins P, Khunti K, et al. Variation in the estimated prevalence of multimorbidity: systematic review and meta-analysis of 193 international studies. BMJ Open. 2022;12(4):e057017-e.

8. Fortin M, Hudon C, Haggerty J, van den Akker M, Almirall J. Prevalence estimates of multimorbidity: a comparative study of two sources. BMC Health Serv Res. 2010;10(1):111-.

9. Fortin M, Almirall J, Nicholson K. Development of a Research Tool to Document Self-Reported Chronic Conditions in Primary Care. J Comorb. 2017;7(1):117–23.

10. Ho ISS, Azoaga-Lorenzo A, Akbari A, Davies J, Khunti K, Kadam U, et al Measuring multimorbidity in research: a Delphi consensus study. BMJ Medicine. July 2022.

11. Herrett E, Gallagher AM, Bhaskaran K, Forbes H, Mathur R, van Staa T, et al. Data Resource Profile: Clinical Practice Research Datalink (CPRD). Int J Epidemiol. 2015;44(3):827–36.

12. The English Indices of Deprivation 2019; Ministry of Housing, Communities and Local Goverment. Available at https://assets.publishing.service.gov.uk/government/uploads/system/uploads/attachment_data/file/853811/IoD2019_FAQ_v4.pdf [accessed 27 March 2022]

13. Lewis JD, Bilker WB, Weinstein RB, Strom BL. The relationship between time since registration and measured incidence rates in the General Practice Research Database. Pharmacoepidemiol Drug Saf. 2005;14(7):443–51.

14. Kuan V, Denaxas S, Gonzalez-Izquierdo A, Direk K, Bhatti O, Husain S, et al. A chronological map of 308 physical and mental health conditions from 4 million individuals in the English National Health Service. Lancet Digit health. 2019;1(2):e63–e77.

15. N’Goran AA, Blaser J, Deruaz-Luyet A, Senn N, Frey P, Haller DM, et al. From chronic conditions to relevance in multimorbidity: A four-step study in family medicine. Fam Pract. 2016;33(4):439–44.

16. Barnett K, Mercer SW, Norbury M, Watt G, Wyke S, Guthrie B. Epidemiology of multimorbidity and implications for health care, research, and medical education: a cross-sectional study. Lancet. 2012;380(9836):37-43.

17. Salive ME. Multimorbidity in older adults. Epidemiol Rev. 2013;35(1):75–83.

18. Charlson ME, Pompei P, Ales KL, MacKenzie CR. A new method of classifying prognostic comorbidity in longitudinal studies: Development and validation. J Chronic Dis. 1987;40(5):373–83.

19. Elixhauser A, Steiner C, Harris DR, Coffey RM. Comorbidity Measures for Use with Administrative Data. Medical care. 1998;36(1):8–27.

20. RDocumentation. ageadjust.direct: Age standardization by direct method. Available at https://www.rdocumentation.org/packages/epitools/versions/0.09/topics/ageadjust.direct [accessed 14 June 2022].

21. Benchimol EI, Smeeth L, Guttmann A, Harron K, Moher D, Petersen I, et al. The REporting of studies Conducted using Observational Routinely-collected health Data (RECORD) statement. PLoS Med. 2015;12(10):e1001885-e.

22. R-Core-Team. R: A Language and Environment for Statistical Computing. R Foundation for Statistical Computing. Vienna, Austria. Available from: http://www.R-project.org. [accessed 27 March 2021] [

23. Simard M, Rahme E, Calfat AC, Sirois C. Multimorbidity measures from health administrative data using ICD system codes: A systematic review. Pharmacoepidemiol Drug Saf. 2022;31(1):1–12.

24. Bagley SC, Altman RB. Computing disease incidence, prevalence and comorbidity from electronic medical records. J Biomed Inform. 2016;63:108–11.

25. Paciorek CJ, Singleton RK, Ikeda N, Laxmaiah A, Widyahening IS, Abarca-Gómez L, et al. Worldwide trends in hypertension prevalence and progress in treatment and control from 1990 to 2019: a pooled analysis of 1201 population-representative studies with 104 million participants. Lancet. 2021;398(10304):957–80.

26. Office for National Statistics. Coronavirus and depression in adults in Great Britain. Available at https://www.ons.gov.uk/peoplepopulationandcommunity/wellbeing/datasets/coronavirusanddepressioninadultsingreatbritain [accessed 19 December 2022]

27. MacRae C, Whittaker H, Mukherjee M, Daines L, Morgan A, Iwundu C, et al. Deriving a Standardised Recommended Respiratory Disease Codelist Repository for Future Research. Pragmat Obs Res. 2022;13:1.

28. Quint JK, Müllerova H, DiSantostefano RL, Forbes H, Eaton S, Hurst JR, et al. Validation of chronic obstructive pulmonary disease recording in the Clinical Practice Research Datalink (CPRD-GOLD). BMJ Open. 2014;4(7):e005540-e.

29. Tonelli M, Wiebe N, Fortin M, Guthrie B, Hemmelgarn BR, James MT, et al. Methods for identifying 30 chronic conditions: application to administrative data. BMC Med Inform Decis Mak. 2015;15(1):31-.

